# Immunoglobulin G1 Fc glycosylation as an early hallmark of severe COVID-19

**DOI:** 10.1101/2021.11.18.21266442

**Authors:** Tamas Pongracz, Jan Nouta, Wenjun Wang, Krista. E. van Meijgaarden, Federica Linty, Gestur Vidarsson, Simone A. Joosten, Tom H. M. Ottenhoff, Cornelis H. Hokke, Jutte J. C. de Vries, Sesmu M. Arbous, Anna H. E. Roukens, Manfred Wuhrer, BEAT-COVID groups, COVID-19 groups

**Affiliations:** Center for Proteomics and Metabolomics, Leiden University Medical Center, Leiden, Netherlands; Dept. of Experimental Immunohematology, Sanquin Research, Amsterdam, Netherlands; Landsteiner Laboratory, Amsterdam University Medical Center, Amsterdam, Netherlands; Dept. of Infectious Diseases, Leiden University Medical Center, Leiden, Netherlands; Dept. of Parasitology, Leiden University Medical Center, Leiden, Netherlands; Dept. of Medical Microbiology, Leiden University Medical Center, Leiden, Netherlands; Dept. of Intensive Care, Leiden University Medical Center, Leiden, Netherlands; Dept. of Internal Medicine, Nephrology, Leiden University Medical Center, Leiden, Netherlands; Dept. of Clinical Epidemiology, Leiden University Medical Center, Leiden, Netherlands; Dept. of Clinical Chemistry, Leiden University Medical Center, Leiden, Netherlands; Dept. of Pulmonary Medicine, Leiden University Medical Center, Leiden, Netherlands; Dept. of Immunology, Leiden University Medical Center, Leiden, Netherlands; Dept. of Internal Medicine, Thrombosis and Hemostasis, Leiden University Medical Center, Leiden, Netherlands; Dept. of Biomedical Data Sciences, Leiden University Medical Center, Leiden, Netherlands; Dept. of Analytical Biosciences, Leiden Academic Centre for Drug Research, Leiden, Netherlands; Dept. of Hematology, Leiden University Medical Center, Leiden, Netherlands; Dept. of Human Genetics, Leiden University Medical Center, Leiden, Netherlands

**Keywords:** IgG glycosylation, anti-Spike IgG, SARS-CoV-2, COVID-19, coronavirus

## Abstract

**Background:** Immunoglobulin G1 (IgG1) effector functions are impacted by the structure of fragment crystallizable (Fc) tail-linked *N*-glycans. Low fucosylation levels on severe acute respiratory syndrome coronavirus 2 (SARS-CoV-2) spike protein specific (anti-S) IgG1 has been described as a hallmark of severe coronavirus disease 2019 (COVID-19) and may lead to activation of macrophages via immune complexes thereby promoting inflammatory responses, altogether suggesting involvement of IgG1 Fc glycosylation modulated immune mechanisms in COVID-19.

**Methods:** In this prospective, observational single center cohort study, IgG1 Fc glycosylation was analyzed by liquid chromatography – mass spectrometry following affinity capturing from serial plasma samples of 159 SARS-CoV-2 infected patients.

**Findings:** At baseline close to disease onset, anti-S IgG1 glycosylation was highly skewed when compared to total plasma IgG1. A rapid, general reduction in glycosylation skewing was observed during the disease course. Low anti-S IgG1 galactosylation and sialylation as well as high bisection were early hallmarks of disease severity, whilst high galactosylation and sialylation and low bisection were found in patients with low disease severity. In line with these observations, anti-S IgG1 glycosylation correlated with various inflammatory markers.

**Interpretation:** Association of low galactosylation, sialylation as well as high bisection with disease severity suggests that Fc-glycan modulated interactions contribute to disease mechanism. Further studies are needed to understand how anti-S IgG1 glycosylation may contributes to disease mechanism and to evaluate its biomarker potential.

**Funding:** This project received funding from the European Commission’s Horizon2020 research and innovation program for H2020-MSCA-ITN IMforFUTURE, under grant agreement number 721815.

**Research in context:** *Evidence before this study:* Antibody glycosylation against the spike (S) protein of patients infected with severe acute respiratory syndrome SARS-CoV-2 has been reported as a potentially important determinant of COVID-19 disease severity. Studies have hitherto focused on afucosylation, a modification on immunoglobulin G1 (IgG) Fc-tail-linked *N*-glycans that enhances effector functions. Most of these studies featured limited sample numbers or were imperfectly matched with respect to demographic and other important confounding factors. Our lab has contributed to some of these studies, and we additionally searched for research articles on PubMed and Google Scholar from January 2020 to October 2021. To date, only two groups studied anti-S IgG1 glycosylation, which resulted in overall three publications found. However, none of these groups found a severity marker between hospitalized non-ICU and ICU patients or studied dynamic changes. Instead, exclusively fucosylation at the first available timepoint has been associated with disease severity between severely ill inpatients and mild outpatients.

*Added value of this study:* In this prospective, observational single center cohort study, we investigated the severity marker potential of anti-S IgG1 glycosylation in severe and mild hospitalized COVID-19 cases, and correlated these findings with numerous inflammation and clinical markers. Our study reveals low galactosylation and sialylation as well as high bisection on anti-S IgG1 as early hallmarks of severe COVID-19, after correction for age and sex effects. In line with these observations, anti-S IgG1 glycosylation correlated with many inflammatory markers. As days since onset is one of the major confounders of anti-S IgG1 glycosylation due to its highly dynamic nature, we additionally confirmed our findings in time-matched patient subgroups. We believe anti-S IgG1 glycosylation may be applicable for patient stratification upon hospitalization.

*Implications of all the available evidence:* Demographic factors as well as temporal differences should be taken into consideration when analyzing IgG1 glycosylation of COVID-19 patients. Anti-S IgG1 glycosylation is highly dynamic, but is a promising early severity marker in COVID-19.

## 1 Introduction

The current global coronavirus disease 19 (COVID-19) pandemic caused by the novel coronavirus severe acute respiratory syndrome coronavirus 2 (SARS-CoV-2) has been leading to extensive hospitalizations worldwide.^1^ To date, more than 253 million infections and more than 5 million deaths have been reported.^2^ SARS-CoV-2 is an enveloped virus and its uptake by target cells in the respiratory tract is mediated by the spike glycoprotein.^1^ Interestingly, most infected people clear the virus with mild symptoms, whilst around 20% of the adult cases are characterized by severe, sometimes life-threatening conditions.^3^ Approximately 7-10 days after symptom onset, seroconversion occurs with immunoglobulin M (IgM) and A (IgA), and G (IgG) antibodies against the spike protein.^4^ These antibodies can form immune complexes with viral particles and thereby neutralize the virus and mediate clearance, but are also capable of aggravating the disease.^5-7^

IgG exerts effector functions via the activation of complement or fragment crystallizable (Fc) gamma receptors (FcγR) on immune cells.^8^ Various effector functions of IgG are steered by the *N*-glycan moiety attached to the highly conserved N297 glycosylation sites within both C_H_2 domains of the Fc tail.^9,10^ Specifically, afucosylated IgG1 shows increased affinity to the activating FcγRIIIa receptor, hence leading to enhanced antibody-dependent cellular cytotoxicity (ADCC).^10,11^ Galactosylated IgG1 shows increased hexamerization, C1q binding and complement activation.^12^

Recent reports have indicated that the high inter-individual variability in COVID-19 disease severity^3^ may partly be explained by low Fc fucosylation of anti-SARS-CoV-2 spike protein-specific (anti-S) IgG1. The lack of core fucose on these specific antibodies early on during disease points to their potential proinflammatory role in severe illness.^6,13,14^ Literature suggests, that in particular membrane-embedded foreign antigens, such as the SARS-CoV-2 spike protein, induce low fucosylated IgG1 responses, which in combination with high titers may lead to excessive macrophage activation and drive COVID-19 associated pathology including acute respiratory distress syndrome.^6,13^

Here, we study the dynamics of IgG1 Fc glycosylation and its association with clinical parameters in a longitudinal cohort of 159 hospitalized COVID-19 patients, analyzing a total of 1300 longitudinal patient samples. We report on the association of early anti-S IgG1 glycosylation signatures with disease severity and various inflammatory markers, indicating its biomarker potential.

## 2 Methods

### 2.1 Chemicals, reagents and enzymes

Type I Ultrapure Water was produced by an ELGA Purelab Ultra system (Elga LabWater, High Wycombe, United Kingdom) and used to create solutions throughout. Ammonium bicarbonate, potassium chloride, formic acid, tolylsulfonyl phenylalanyl chloromethyl ketone-treated trypsin from bovine pancreas was obtained from Sigma-Aldrich (Steinheim, Germany). Trifluoroacetic acid, disodium hydrogen phosphate dihydrate, potassium dihydrogen phosphate, and sodium chloride were purchased from Merck (Darmstadt, Germany). HPLC-supra-gradient acetonitrile was obtained from Biosolve (Valkenswaard, The Netherlands). The Visucon-F pooled healthy human plasma standard originated from Affinity Biologicals (Ancaster, Canada). Protein G Sepharose 4 Fast Flow beads were obtained from GE Healthcare (Uppsala, Sweden). Recombinant trimerized spike protein was prepared as described.^15^

### 2.2 Study cohort

BEAT-COVID-19 is a prospective, observational single center cohort study established at Leiden University Medical Center, with longitudinal plasma samples of 159 PCR-confirmed SARS-CoV-2 infected hospitalized patients that were collected during the first and second wave of the pandemic (between May 2020 and October 2020) **(Table 1, Table S1, Figure S1)**. After informed consent was obtained from the patient or his/her relatives, longitudinal sampling was performed for the duration of the hospital admission, and one convalescent sample was obtained at the outpatient follow-up appointment, which was scheduled six weeks after hospital discharge. Statistical sample size calculation was not performed, the sample size was determined based on availability. The Medical Ethics Committee Leiden-Den Haag-Delft (NL73740.058.20) approved the study. The trial was registered in the Dutch Trial Registry (NL8589). The study complied with the latest version of the Declaration of Helsinki.

**Table 1.** Baseline patient characteristics. Median and interquartile ranges are shown unless indicated otherwise. The sex of one patient is unknown (not shown).

|  | ICU (n=77) | non-ICU (n=82) |
| --- | --- | --- |
| Age | 65 (59-71) | 66.5 (54-74.5) |
| Female, n (%) | 18 (23) | 21 (26) |
| Male, n (%) | 59 (77) | 60 (74) |
| Severity score | 12 (10-14) | 3 (2-4) |
| Days since symptom onset | 15.5 (12-22) | 13 (10-16) |

### 2.3 Sample preparation for IgG Fc glycosylation analysis

Anti-S IgG was captured using a setup that resembles a conventional ELISA: IgGs were affinity-captured from plasma using recombinant trimerized spike-protein-coated Maxisorp NUNC-Immuno plate (Thermo Fisher Scientific, Roskilde, Denmark), whereas total IgG was affinity-captured using protein G Sepharose Fast Flow 4 beads, as described previously.^13,16^ Antibodies were eluted using 100 mM formic acid and the samples were dried by vacuum centrifugation. Samples were reconstituted in 25 mM ammonium bicarbonate and subjected to tryptic cleavage, as described elsewhere.^16^ Samples belonging to a single patient were prepared and measured consecutively on the same plate, except for follow-up samples after hospitalization period. On each plate, at least 3 Visucon-F plasma standards (dating pre-COVID-19) and 3 blanks were included.

### 2.4 IgG Fc glycosylation analysis

Glycopeptides were separated and detected using an Ultimate 3000 high-performance liquid chromatography (HPLC) system (Thermo Fisher Scientific, Waltham, MA) hyphenated to an Impact quadrupole time-of-flight mass spectrometer (Bruker Daltonics, Billerica, MA), as described.^16^

### 2.5 Liquid chromatograph – mass spectrometry data processing

MzXML files were generated from raw liquid chromatograph – mass spectrometry (LC-MS) spectra. An in-house developed software, LaCyTools was used for the alignment and targeted extraction of raw data.^17^ Alignment was performed based on the average retention time of minimum three abundant IgG1 glycoforms. The targeted extraction list included analytes of the 2^+^ and 3^+^ charge states and was based on manual annotation of the mass spectra as well as on literature.^18,19^ A pre-COVID-19 plasma pool (Visucon-F) was measured in triplicate in each plate to assess method robustness and was as well used as negative control. All spectra below the average intensity plus three times the standard deviation of negative controls was excluded from further analysis. Signals were integrated by covering a minimum of 95% of the area of the isotopic envelope of glycopeptide peaks. Inclusion of an analyte for the final data analysis was based on quality criteria such as signal-to-noise (> 9), isotopic pattern quality (< 25% deviation from the theoretical isotopic pattern), and mass error (within a ± 20 ppm range). Furthermore, analytes that were present in at least 1 out of 4 anti-S IgG1 spectra (25%) were included in the final analysis.

### 2.6 Cytokine measurements by cytometric bead array

Circulating cytokine and chemokine levels were determined in serum using commercially available bead based multiplex assays using the BioPLex 100 system for acquisition as previously described.^20^ Standard curves were included in the kits and, in addition, a pooled serum sample of 4 hospital admitted COVID-19 patients was included as internal reference in all assays. Four commercially available kits were used: Bio-Plex Pro™ Human Cytokine Screening Panel 48-plex, Bio-Plex Pro^tm^ Human Chemokine Panel 40-Plex, Bio-Plex Pro^tm^ Human Inflammation Panel 1 and 37-Plex; Bio-Plex Pro^tm^ Human Th17 panel (IL-17F, IL-21, IL-23, IL-25, IL-31, IL-33) (all obtained from Bio-Rad, Veenendaal, The Netherlands).

### 2.7 Antibody titer measurement

Semi-quantitative detection of SARS-CoV-2 anti-nucleocapsid (N) protein IgG was performed on the Abbott Architect platform.^21-23^ In this antibody chemiluminescent microparticle immunoassay (CMIA) test, the SARS-CoV-2 antigen coated paramagnetic microparticles bind to the IgG antibodies that attach to the viral nucleocapsid protein in human serum samples. The Sample/Calibrator index values of chemiluminescence in relative light units (RLU) of 1·40 (IgG assay) respectively 1·00 (IgM assay) and above were considered as positive per the manufacturer’s instructions.

Quantitative detection of SARS-CoV-2 anti-S1/S2 IgG antibodies was performed using the DiaSorin LIAISON platform. The CLIA assay consists of paramagnetic microparticles coated with distally biotinylated S1 and S2 fragments of the viral surface spike protein. RLUs proportional to the sample’s anti-S1/S2 IgG levels are converted to AU/mL based on a standardized master curve.

Semi-quantitative detection of SARS-CoV-2 anti-RBD IgM antibodies was performed using the Wantai IgM-ELISA (CE-IVD) kit (Sanbio).^24^ Briefly, the IgM u-chain capture method was used to detect IgM antibodies using a double-antigen sandwich immunoassay using mammalian cell-expressed recombinant antigens containing the RBD of the spike protein of SARS-CoV-2 and the immobilized and horseradish peroxidase-conjugated antigen. Sample/Cut-off index OD values of 1 and higher were considered positive per the manufacturer’s instructions.

Semi-quantitative detection of SARS-CoV-2 anti-S1 IgA antibodies was performed using the Euroimmun IgA 2-step ELISA.^25^ Ratio values of 1·1 and higher were considered positive per the manufacturer’s instructions.

### 2.8 Severity score calculation

The severity score is based on the 4C mortality score.^26^ The 4C mortality score is a prediction score calculated at admission, and the severity score calculated in our cohort represents the daily clinical disease severity, and thus is dependent on parameters that can change over time. Therefore, the fixed parameters of the 4C score were removed (i.e. age, sex at birth, number of comorbidities. Daily oxygen flow for non-ICU patients (L/min) and p/f ratio (kPa) and FiO2 (%) for ICU patients were added to our severity score **(Table S2)**.

### 2.9 Statistical analysis

Relative intensity of each glycopeptide species in the final analyte list was calculated by normalizing to the sum of their total areas **(Table S3)**. Structurally similar glycopeptide species were used for the calculation of derived traits fucosylation, bisection, galactosylation and sialylation **(Table S4)**. Anti-S and total IgG1 glycosylation traits were compared using a Wilcoxon signed-rank test **(Figure 1, Table S5)**, while a Wilcoxon rank-sum test was used to compare non-ICU and ICU patients as well as severity score groups (**Figure 3-4; Table S6-7; Figure S3, Figure S9-10**). To account for multiple testing, *p*-values of the Wilcoxon-tests have been corrected by the Benjamini-Hochberg procedure to 5% FDR in each statistical question **(Table S5-7)**. Spearman’s correlation was used to explore associations between glycosylation traits and age **(Figure S2)**, as well as between glycosylation traits and inflammatory markers and titers **(Figure 5, Table S8)**. To assess method repeatability, the inter-plate variation for the 14 analytes included in the final analysis was calculated for the standards, which was 2·4%. All statistical analyses and visualizations were performed in R, version 4.1.0 (R Foundation for Statistical Computing, Vienna, Austria) and RStudio, version 1.4.1717 (RStudio, Boston, MA).

**Figure 1.**
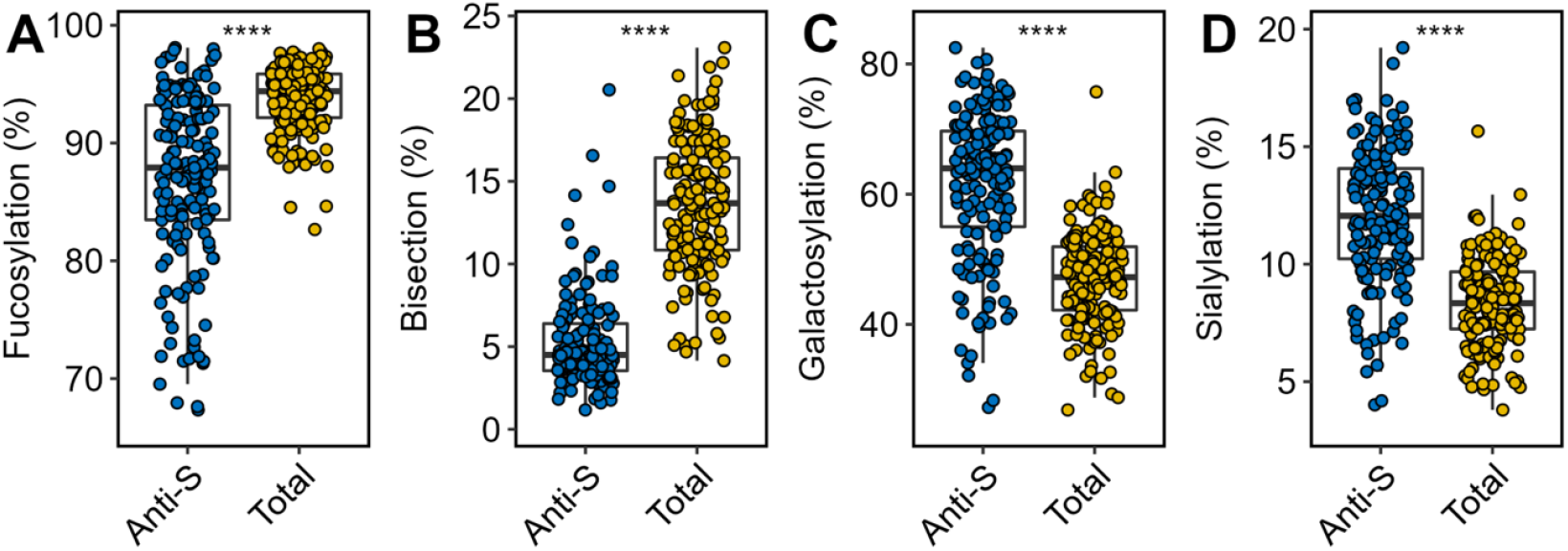
Comparison of anti-S (blue) and total (yellow) IgG1 Fc glycosylation. Relative abundance of IgG1 **(A)** fucosylation, **(B)** bisection, **(C)** galactosylation and **(D)** sialylation of anti-S and total IgG1 are given at hospitalization (n=159). Boxplots display the median and the interquartile range, whereas whiskers represent the first and third quartiles. A Wilcoxon signed-rank test was used to compare anti-S with total IgG1. ****: *p*-value < 0·0001.

**Figure 2.**
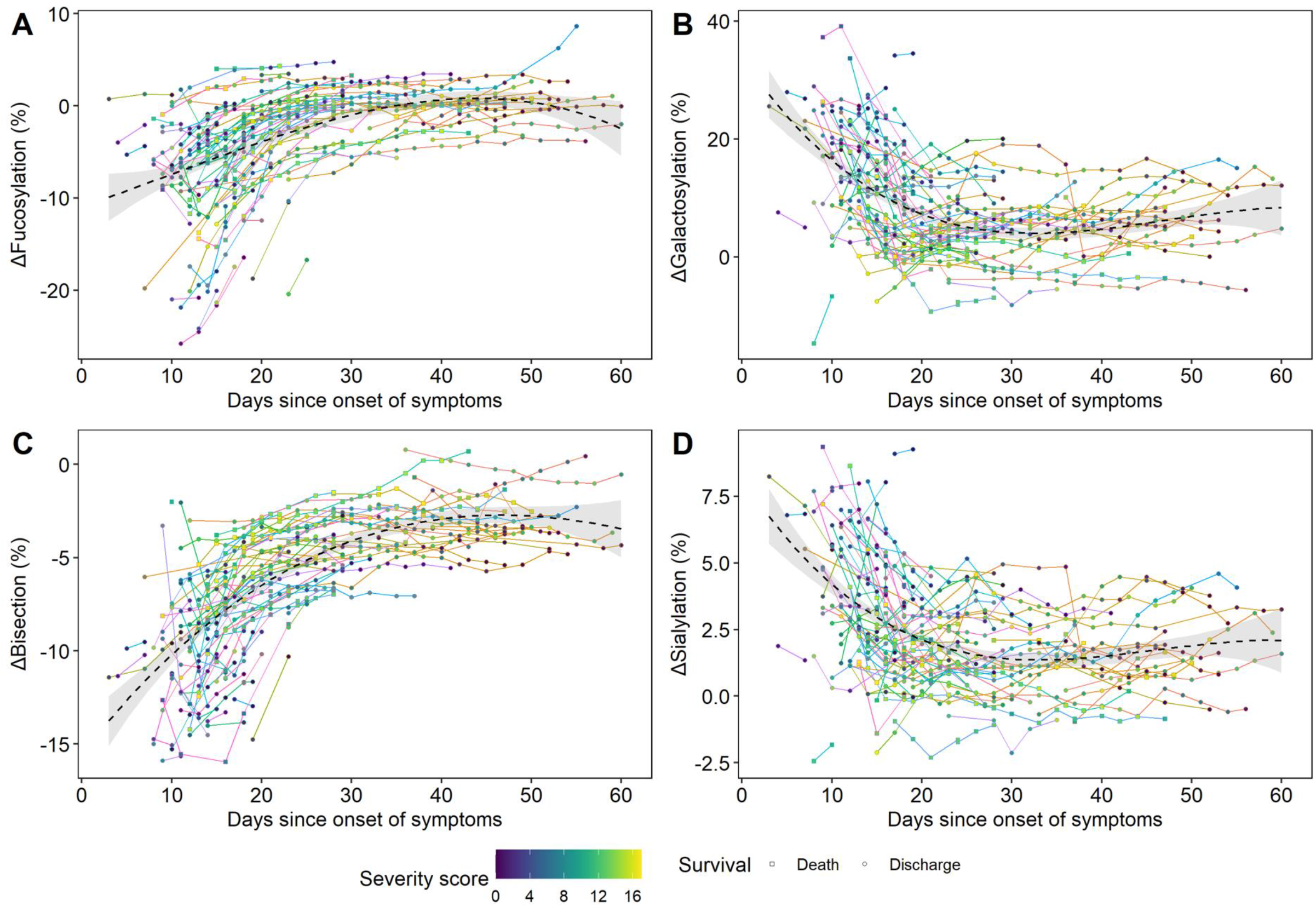
ΔGlycosylation dynamics until 60 days since symptom onset. The time course of Δglycosylation traits **(A)** fucosylation, **(B)** galactosylation, **(C)** bisection and **(D)** sialylation as shown during the hospitalization period (n=109). Line colors correspond to a single COVID-19 patient, whilst the color gradient in the circles/squares indicate the corresponding severity score (grey = NA). The shape displays whether a patient passed away (square) or was discharged alive (circle). The black dashed line with a grey 95% confidence interval band is a cubic polynomial fit over the shown datapoints to illustrate overall dynamics. Late timepoints and two outliers are shown in the **Supplementary Material** due to spatial constraints **(Figure S4-5)**, as well as anti-S and total IgG1 glycosylation dynamics **(Figure S6)**.

**Figure 3.**
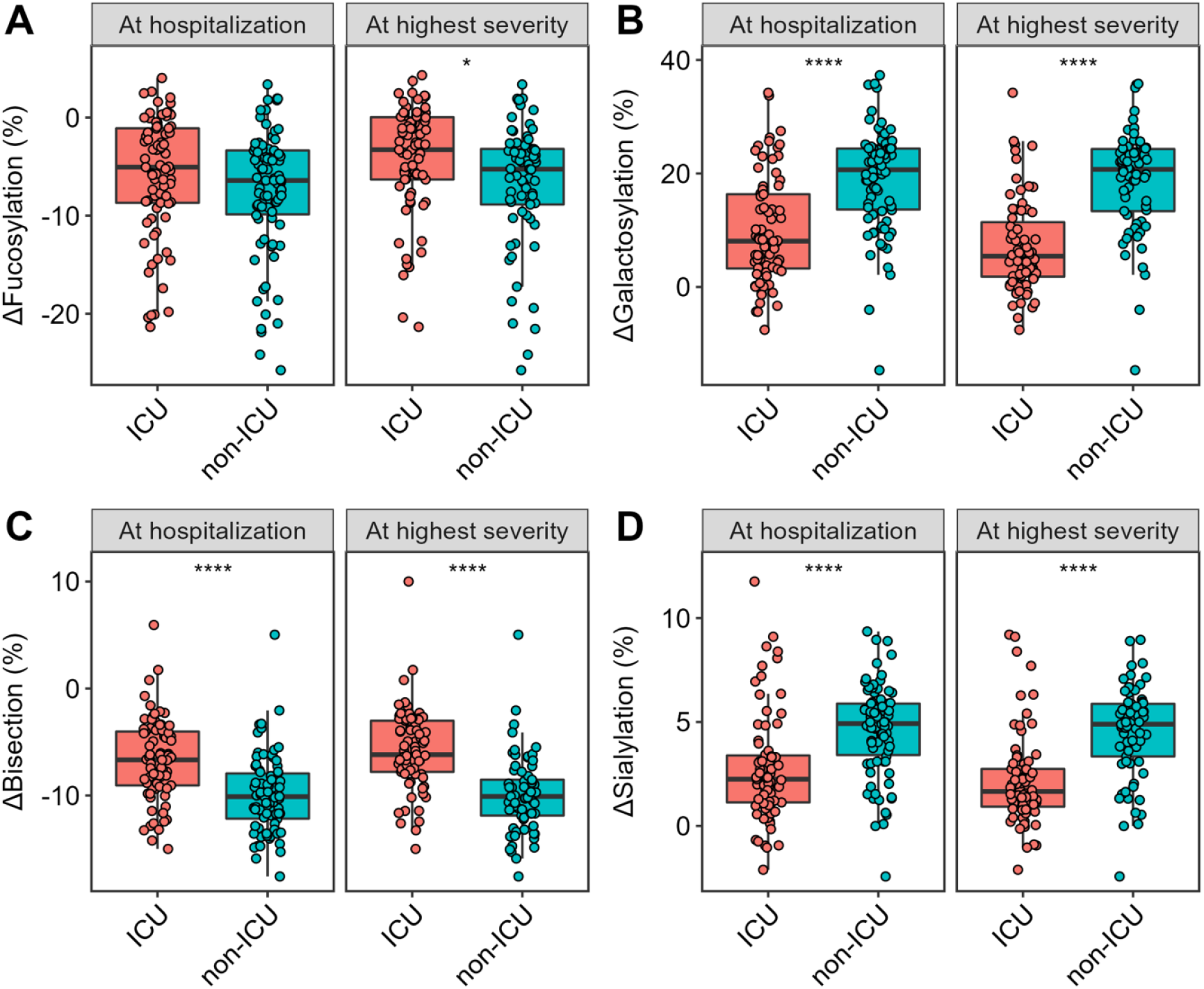
Comparison of Δglycosylation traits of patients admitted to ICU (red) or non-ICU (blue) treatment. Shown in the facets are the relative levels of ΔIgG1 **(A)** fucosylation, **(B)** galactosylation, **(C)** bisection and **(D)** sialylation at the time of hospitalization (left; n=159; 77 ICU and 82 non-ICU patients, respectively) and at the time of highest disease severity (right; n=144; 75 ICU and 69 non-ICU patients, respectively). The highest severity timepoint has been defined for each patient as the earliest possible timepoint with highest severity score during hospitalization. A Wilcoxon rank-sum test was used to compare ICU and non-ICU patients **(Table S6)**. *, ****: *p*-value < 0·05, 0·0001, respectively. Glycosylation dynamics of ICU and non-ICU patients between day 10 and 25 are shown in **Figure S8**.

## 3 Results

Both anti-S and total IgG1 glycosylation signature of 159 COVID-19 patients (39 female and 119 male) and corresponding timepoints were analyzed during their entire hospitalization period. The patient demographics and the comprehensive cohort characteristics are presented in **Table 1** and **Table S1**, respectively. Follow-up samples after hospital discharge were available for 19 patients **(Table S1, Figure S5)**. LC-MS was employed to analyze Fc glycosylation on the glycopeptide level after tryptic digestion, which allowed the identification of 14 glycoforms. The found glycoforms were consistent with previous reports on anti-S IgG1 glycosylation^13,14^, from which fucosylation, bisection, galactosylation and sialylation were calculated **(Table S3-4)**. Overall, a total of 650 total IgG1 and 650 anti-S IgG1 glycosylation profiles were determined.

### 3.1 Anti-S IgG1 Fc glycosylation of COVID-19 patients is skewed

The Fc glycosylation signatures of anti-S and total IgG1 were compared pairwise at hospitalization with regard to fucosylation, bisection, galactosylation and sialylation **(Figure 1, Table S5)**. Fucosylation of anti-S was significantly lower than total (fold change (FC): 0·93; *p*-value: 3·4×10^−24^) **(Figure 1A, Table S5)**. Notably, a prominently low anti-S fucosylation (<85%) was found for 56 patients, with a few patients showing levels as low as 66% **(Figure 1A)**. Similarly, bisection of anti-S was markedly lower than total IgG1 (FC: 0·33; *p*-value: 3·1×10^−27^) **(Figure 1B)**. Anti-S galactosylation (FC: 1·35 ; *p*-value: 8·1×10^−26^) **(Figure 1C)** and sialylation (FC: 1·45; *p*-value: 2·7×10^−26^) **(Figure 1D)** were elevated as compared to their total IgG1 counterpart.

### 3.2 Dynamic regulation of IgG1 Fc glycosylation in COVID-19

Next, we explored the changes of glycosylation over time. Anti-S glycosylation was found to be highly dynamic, but also total IgG1 glycosylation showed changes in the course of the disease **(Figure S6**). Both anti-S and total IgG1 galactosylation was found to be confounded by age and sex **(Figure S2)** in line with literature on IgG Fc glycosylation.^27^ Therefore, delta (Δ) values were calculated by subtracting total from anti-S IgG1 levels to eliminate the confounding effect, and used hereafter **(Figure S2-3)**.

The longitudinal samples allowed us to establish the time course of ΔIgG1 glycosylation during hospitalization, normalized for day of onset of symptoms **(Figure 2, Table S1)**. Interestingly, all glycosylation traits showed a transient pattern for most patients, and were characterized by profound dynamics, as illustrated by the timelines of individual patients (as indicated by differential line coloring) and by the fit cubic polynomial line **(Figure 2)**. Fucosylation **(Figure 2A)** and bisection **(Figure 2C)** showed a rapid increase within days and weeks after onset of the disease, followed by a plateau and approximation of the glycosylation patterns of total IgG1 **(Figure S6)**. In contrast, galactosylation **(Figure 2B)** and sialylation **(Figure 2D)** quickly declined in the first days and weeks, with the decrease continuing for a long period albeit at lower pace. At the moment of hospital discharge anti-S galactosylation and sialylation were still slightly higher than total IgG1. Since 19 patients returned for follow-up sampling after hospital discharge, we noted that for most, fucosylation and bisection largely remained constant or slightly increased, whilst galactosylation and sialylation continued to decrease since the last available timepoint **(Figure S5)**.

### 3.3 IgG1 Fc glycosylation associates with ICU admission

To investigate whether Fc glycosylation was associated with intensive care unit (ICU) admission, patients were stratified based on treatment need. This resulted in two groups: 1) patients who at some point during hospitalization were admitted to the ICU, and 2) patients who were not enrolled to ICU treatment at all (non-ICU) during hospitalization. ΔIgG1 glycosylation derived traits fucosylation, bisection, galactosylation and sialylation of the above groups were compared both at time of hospitalization and at the time point of their highest disease severity **(Figure 3, Table S6)**.

ΔIgG1 Fc glycosylation of ICU patients showed a different profile from those of non-ICU patients, with the latter being characterized by lower bisection (FC: 0·66, *p*-value: 7·2×10^−8^) **(Figure 3C)**, and higher galactosylation (FC: 0·39, *p*-value: 2·9×10^−9^) **(Figure 3B)** and sialylation (FC: 0·46, *p*-value: 1·7×10^−7^) at the time of hospitalization **(Figure 3D)**. This difference was maintained or even more pronounced at the time of highest disease severity (FC: 0·61, 0·26, 0·34; *p*-value: 1·9×10^−10^, 4·1×10^−12^, 3·4×10^−9^, for Δbisection, Δgalactosylation and Δsialylation, respectively) **(Table S6)**. Fucosylation levels of the ICU group were higher at the time of highest disease severity (FC 0·62; *p*-value: 0·012), but remained similar at the time of hospital admission **(Figure 3A)**. To confirm that the observed effects were not confounded by vast glycosylation dynamics, a subset of non-ICU and ICU patients were created and compared, which resulted in comparable observations with regards to Δbisection, Δgalactosylation and Δsialylation as shown above **(Figure S7-8)**.

### 3.4 IgG1 Fc glycosylation associates with disease severity

Patients were stratified into three groups based on their severity score: 1) severity score between 0-5 (low severity), 2) 6-11 (intermediate severity) and 3) 12-17 (high severity). Similarly as before, ΔIgG1 glycosylation traits were compared both at time of hospitalization and at time of highest disease severity **(Figure 4, Table S7)**.

**Figure 4.**
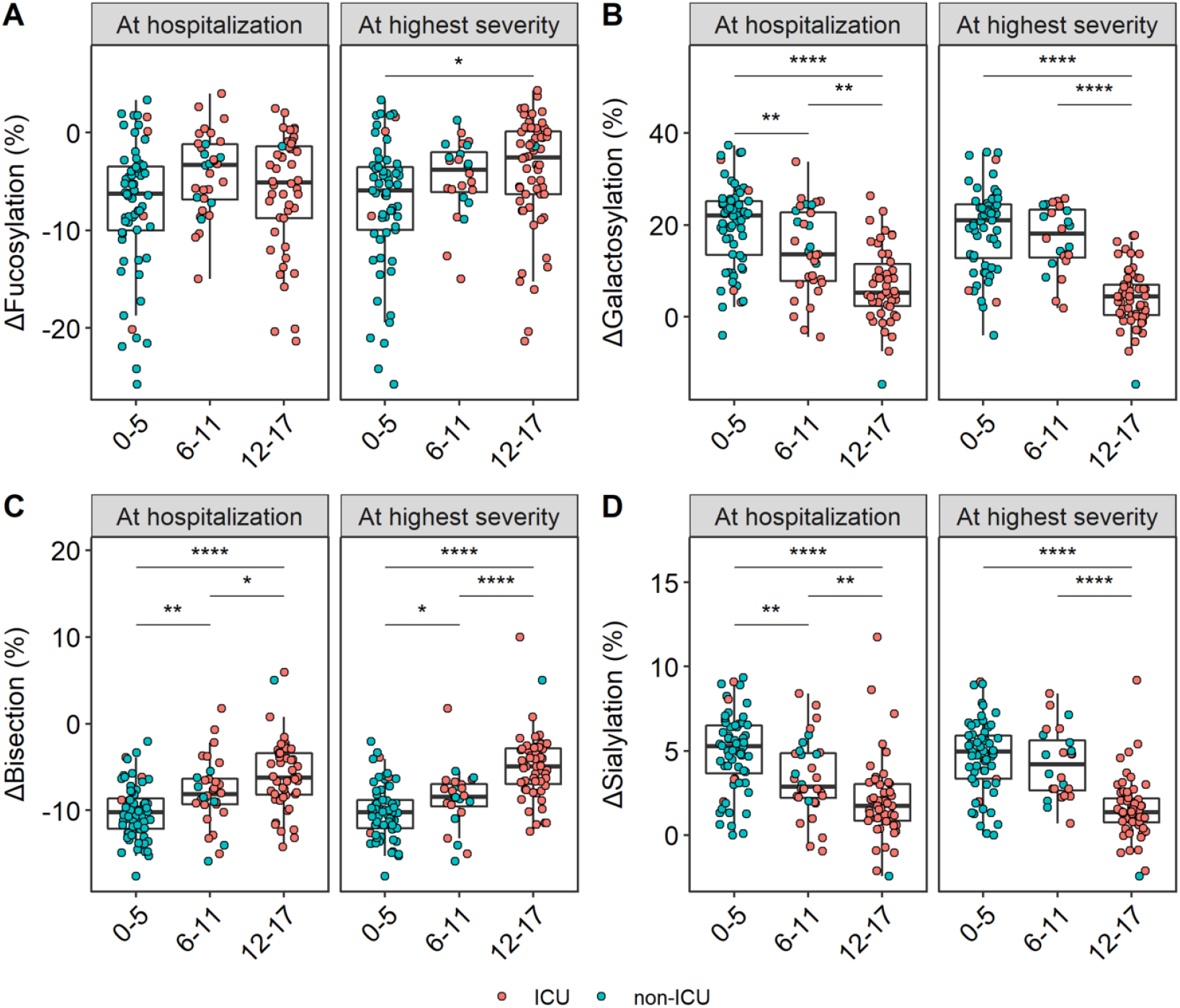
Comparison of Δglycosylation of patients in different severity score groups. Shown in the facets are the relative levels of ΔIgG1 **(A)** fucosylation, **(B)** bisection, **(C)** galactosylation and **(D)** sialylation at the time of hospitalization (left; n=142; 64 low severity, 32 intermediate severity and 46 high severity patients, respectively) and at the time of highest disease severity (right; n=144 n=144; 61 low severity, 24 intermediate severity and 59 high severity patients, respectively). Color indicates ICU (red) and non-ICU (blue) patients. A Wilcoxon rank-sum test was used to compare the different severity score groups **(Table S7)**. *, **, ****: *p*-value < 0·05, 0·01, 0·0001, respectively.

ΔBisection was found to be increased in groups with increased disease severity **(Figure 4C)**, whereas Δgalactosylation **(Figure 4B)** and Δsialylation **(Figure 4D)** patterns were found to be decreased with increased disease severity at the time of hospitalization **(Table S7)**. These observations were largely maintained at highest disease severity **(Figure 4, Table S7)**. Higher fucosylation marked the time of highest disease severity, but remained rather stable at the time of hospital admission between all groups **(Figure 4A, Table S7)**. To confirm that the observed effects were not confounded due to profound glycosylation dynamics, subsets of patients matched for the time since disease onset were compared, which resulted in similar observations with regards to Δgalactosylation and Δsialylation as shown above, whereas we could not exclude a potential confounding effect for the bisection signature, maybe caused by swift glycosylation dynamics, low sample size, or the combination thereof **(Figure S9)**. Apart from ICU admission and severity score, we tested acute respiratory syndrome, ventilation and survival, and found Δbisection being higher for patients at baseline who passed away later **(Figure S10)**.

### 3.5 IgG1 Fc glycosylation associates with inflammatory markers

Multiple inflammatory mediators (in serum) and clinical parameters were measured for patients enrolled during the first wave of the pandemic. These include members of the CXC, CCL and CX3C chemokine families, cytokines and corresponding soluble receptors, acute phase proteins and other mediators involved in the immune response as well as severity scores and anti-viral antibody titers. In general, negative associations were found between Δgalactosylation and Δsialylation and positive associations for Δbisection and Δfucosylation with inflammatory markers at baseline. One notable exception was a strong negative correlation between anti-RBD IgM levels and Δbisection and Δfucosylation at baseline and at highest severity, respectively. ΔSialylation associated negatively with various chemokines, such as CCL24 (r = -0·45), CX3CL1 (r = -0·43), CCL25 (r = -0·34), certain cytokines, such as IL-8 (r = -0·29), IFN-γ (r = -0·3) and several other variables **(Figure 5, Table S8)**.

**Figure 5.**
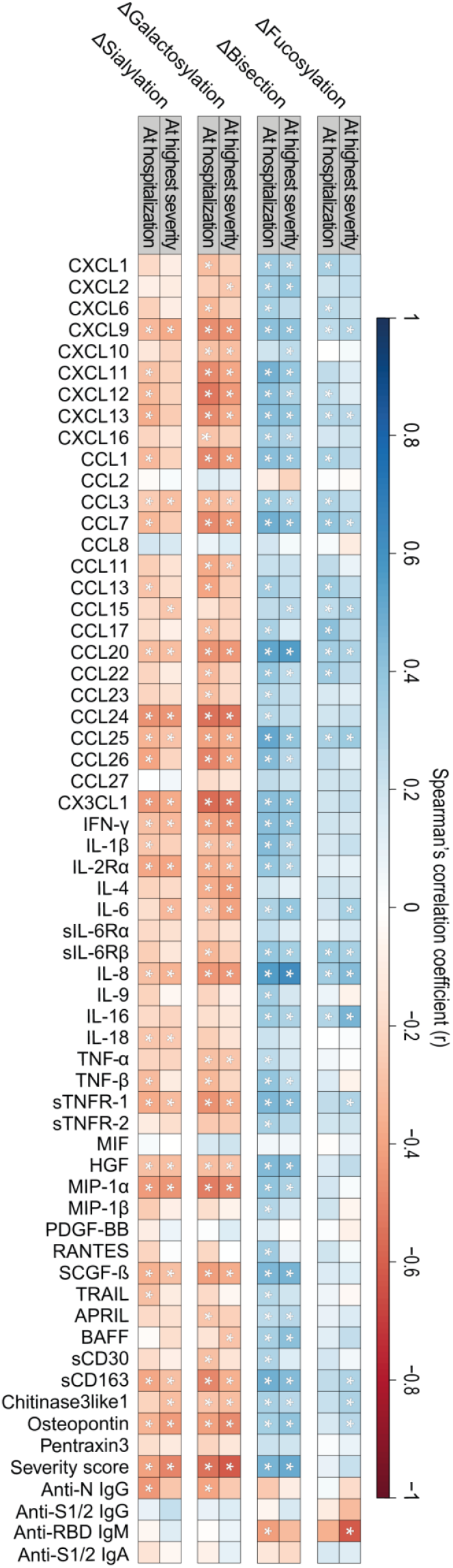
Heatmap visualizing Spearman’s correlations between Δglycosylation traits and inflammatory markers at time of hospitalization (left side of each panel; n=58) and at time of highest disease severity (right side of each panel; n=59). Asterisk (*) indicates a significant Spearman’s correlation (*p*-value < 0.05).

Comparable, and largely overlapping negative associations were found for Δgalactosylation as for Δsialylation: CCL24 (r = -0·55), CX3CL1 (r = -0·56), CCL25 (r = -0·41), IL-8 (r = -0·44), INF-γ (r = -0·4) and TNF-β (r = -0·33). Conversely, Δbisection associated positively with IL-8 (r = 0·56), CCL25 (r = 0·52) and CX3CL1 (r = 0·56). Additionally, severity score negatively correlated with Δgalactosylation (r = -0·55) and Δsialylation (r = -0·41) and positively with Δbisection (r = 0·46). Positive associations were found between Δfucosylation and inflammatory markers, including CCL17 (r = 0·41) and IL-8 (r = 0·34). The above described baseline correlations were comparable to those at the time of highest disease severity, but a vast body of associations were temporary **(Figure 5, Table S8)**.

## 4 Discussion

In this study, we analyzed total and anti-S IgG1 Fc glycosylation of 159 COVID-19 patients at different timepoints during their clinical illness. Although several studies reported on the importance of (anti-S) IgG1 Fc glycosylation and its association with disease severity in COVID-19^6,13,14,28,29^, this study involves a large, single center cohort that confirms specific anti-S IgG1 glycosylation features as an early hallmark of severe COVID-19 in an age-and sex-corrected, time-matched dataset at baseline, and in the longitudinal dimension.

Afucosylated IgG1 B cell responses have recently been described to characterize immune reactions against membrane-embedded antigens in general, and in particular against viral infections caused by enveloped viruses such as COVID-19.^13^ Foregoing studies showed that severe, hospitalized patients exhibit a decreased anti-S IgG1 fucosylation as compared to mild, non-hospitalized patients.^6,13,14^ Accordingly, we likewise observed proinflammatory, low-fucosylation signatures on anti-S as compared to total IgG1, but found no difference in fucosylation comparing hospitalized ICU patients versus hospitalized non-ICU patients, which is in line with a previous report on anti-SARS-CoV-2 receptor binding domain (anti-RBD) IgG1 fucosylation.^14^ Therefore, based on the early existence of these proinflammatory signatures in some of the patients, we hypothesize that low fucosylation – potentially even lower before measurable seroconversion, as hypothesized before^13^ – on anti-S IgG1 may act as an early inflammatory signal that promotes the development of a more severe disease in COVID-19 patients, resulting in hospital admission. However, disease severity between hospitalized patients could not be further distinguished based on anti-S IgG1 fucosylation. Furthermore, hardly any negative associations were found between anti-S IgG1 fucosylation and inflammatory markers in this study, unlike in previous reports, where *in vitro* experiments demonstrated that the stimulation of isolated macrophages with recombinant, glycoengineered anti-S or patient sera-derived low-fucose IgG1 antibodies trigger higher proinflammatory cytokine release than those with normal fucose levels.^6,13,14^ However, high proinflammatory cytokine levels are not necessarily present in all severe patients^30^, and this contrasting observation suggests a different regulation and/or the temporal resolution of fucosylation and cytokine production dynamics *in vivo*. Additionally, beyond or in combination with low anti-S IgG1 fucosylation a pre-existing risk factor may plays a role in COVID-19 disease severity, which hitherto remained unclear.^29^ Of note, the anti-S and anti-RBD IgG1 Fc glycosylation data were all determined from the circulation, and it is unclear to which extent this would reflect the inflammatory pattern and glycosylation profile of anti-S antibodies in the lung. Our results demonstrate that the proinflammatory fucosylation signature that is observed at the early time points in the disease tends to fade with the course of the disease, which one may interpret as a shift towards a more anti-inflammatory Fc glycosylation profile that is maintained over time. The absence of core fucose is known to enhance a proinflammatory immune response by activating FcγRIII receptors on monocytes, macrophages and NK cells.^10^ Decreased fucosylation on specific IgG1 has been described in HIV^13,31^ and dengue fever^32^, as well as in alloimmune diseases.^33-37^ However, whilst afucosylation of specific IgG1 plays a protective role in HIV, it clearly marks high disease severity in dengue, alloimmune diseases or COVID-19.^6,13,14^ Furthermore, low total IgG1 fucosylation has been associated with outcome of pediatric meningococcal sepsis indicating a systemic inflammation due to the potential accumulation of airway infections during early childhood.^38^ Even though the origin of low fucose IgG responses is seemingly linked to antigen context and affect mostly specific antibodies^13^, the mechanisms underlying the dynamics of antibody glycosylation remain elusive.

Besides afucosylation, a transient, decreased bisection was found on anti-S IgG1. Recent reports suggest that severe COVID-19 patients present low levels of bisection both on total IgG (Fc and Fab combined)^29^ and anti-S IgG1^13^ relative to mild cases. In contrast, no difference was found in anti-RBD IgG1 bisection between ICU and non-ICU patients in age- and sex-matched patients^14^, albeit these disease groups were largely comparable to the ones in our study. While bisection associated positively with ICU admission, disease severity and survival in our study, it lacks functional relevance based on our current understanding and has no effect on FcγRIII or C1q binding.^10,39^

Elevated galactosylation and sialylation of anti-S IgG1 were associated with a less severe disease course upon hospitalization, and no ICU admission. Similar observations were made in a previous report, where severe COVID-19 was characterized by lower anti-S IgG1 galactosylation and sialylation than mild COVID-19.^13^ Interestingly, both anti-S and total IgG1 galactosylation and sialylation decrease by advancing age. As Larsen et al. compared anti-S IgG1 galactosylation and sialylation of imperfectly age matched patient groups without age correction, the disease and age effects remained indiscernible.^13^ We describe decreased anti-S IgG1 galactosylation in ICU patients as compared to non-ICU patients, and analogously, markedly lower specific IgG1 galactosylation has been shown to characterize the more severe, active phase of tuberculosis as compared to its latent counterpart.^40^ Even though more and more reports support that elevated levels of galactosylated IgG are associated with the activation of the classical complement pathway^10,12,41^, galactosylation was associated with increased disease severity in this study, possibly due to the fact that complement can contribute to the increased inflammation both directly, and through inducing a chemotactic response through C5a, thereby increasing cellular infiltration to inflamed tissues such as the lung.^42^ Elevated sialylation levels on anti-S IgG1 were associated with increased disease severity in the current report. Sialylation has been broadly described as critical in mediating anti-inflammatory activity^43-45^, yet it remains to be elucidated whether sialylated IgG exerts an anti-inflammatory effect in COVID-19.

## 5 Conclusions

This study established anti-S IgG1 bisection, galactosylation and sialylation as a unique combination of features that associate with ICU admission and disease severity in hospitalized COVID-19 patients. These features were additionally associated with markers of inflammation. Hence, we believe anti-S IgG1 glycosylation may be applicable for patient stratification upon hospitalization. The glycosylation profiles are highly dynamic, the drivers of which remain elusive and to be investigated in future studies.

## Data Availability

All data produced in the present study are available upon reasonable request to the authors.

## 6 Contributors

T. P.: Data (pre)processing, data curation, formal analysis, validation, investigation, visualization, statistical analysis, data interpretation, conceptualization, writing – original draft preparation. J. N.: sample preparation, data acquisition (IgG Fc glycosylation), W. W.: sample preparation (IgG Fc glycosylation), F. L.: production and purification of recombinant spike protein, K. E. van M, S. A. J., T. H. M. O: data acquisition (soluble marker profiles), review J. J. C. V.: data acquisition (antibody titers), writing – review & editing, A. H. E. R., S. M. A.: set up of cohort, recruitment & sampling of participants, writing – review & editing, G.V., C. H. H.: writing – reviewing & editing M. W.: Supervision, writing – review & editing, conceptualization, funding acquisition.

All authors were involved in the critical revision of the manuscript and have given approval to the final version of the manuscript.

## 9 Data sharing

The datasets generated for this study are available on request from the corresponding author.

## Supporting information

**Supplementary Figure 1.**
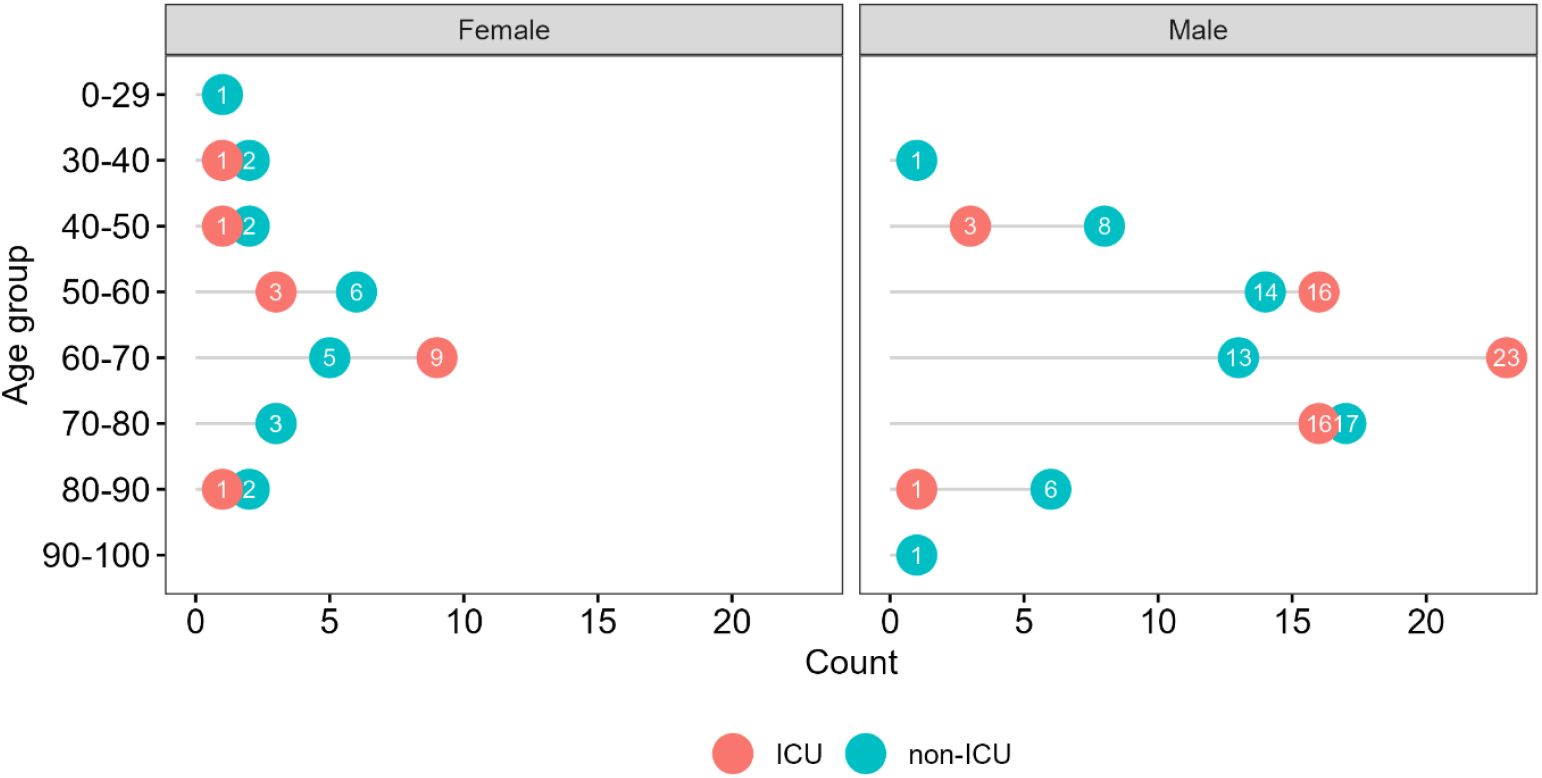
Age and sex distribution in the BEAT-COVID cohort. Overall 159 patients participated in the study (39 female and 119 male, 1 unknown (not shown)). The color illustrates ICU (red) and non-ICU (blue) treatment groups, whereas the number in the circles indicates the number of patients in the corresponding group.

**Supplementary Figure 2.**
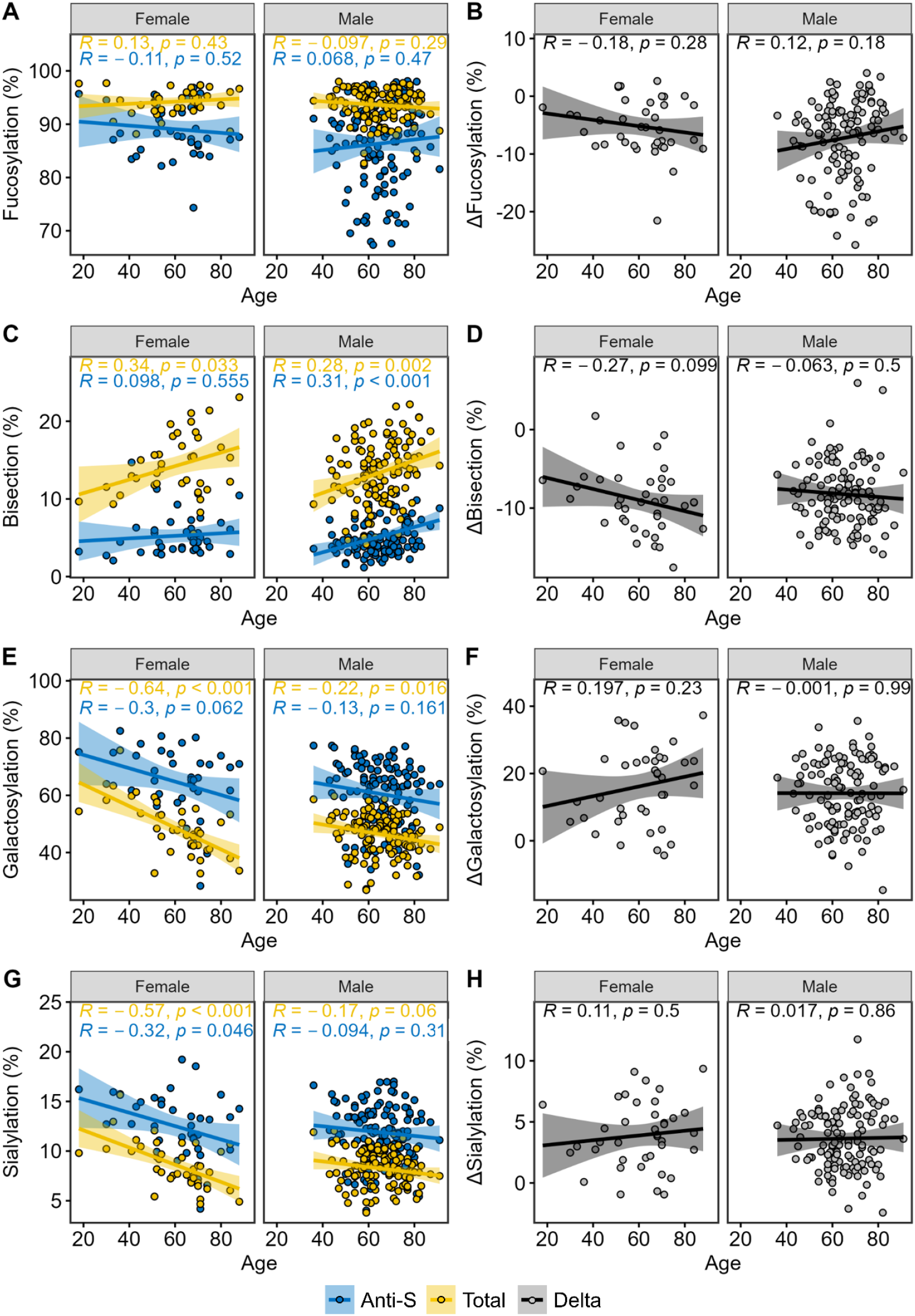
Anti-S (blue) and total (yellow) IgG1 bisection, galactosylation and sialylation are confounded by age in a similar way, which is eliminated by normalizing to total IgG1 levels. IgG1 **(A)** fucosylation, **(C)** bisection, **(E)** galactosylation and **(G)** sialylation as a proxy of age in female (left) and male (right) patients. Corresponding delta (Δ) IgG1 **(B)** fucosylation and **(D)** bisection, **(F)** galactosylation and **(H)** sialylation levels (all in grey), as normalized to total IgG levels by subtracting total from anti-S IgG1 glycosylation levels. Baseline timepoints are shown. Shown in the inset are the Spearmen correlation coefficients (R) and *p*-values, respectively. IgG1 bisection is known to increase, whereas galactosylation and sialylation are known to decrease upon aging.^1^ Correction for the age confounding effect was performed by normalizing to total IgG levels, as illustrated by the weak and non-significant Spearman correlations and *p*-values, respectively **(B, D, F, H)**.

**Supplementary Figure 3.**
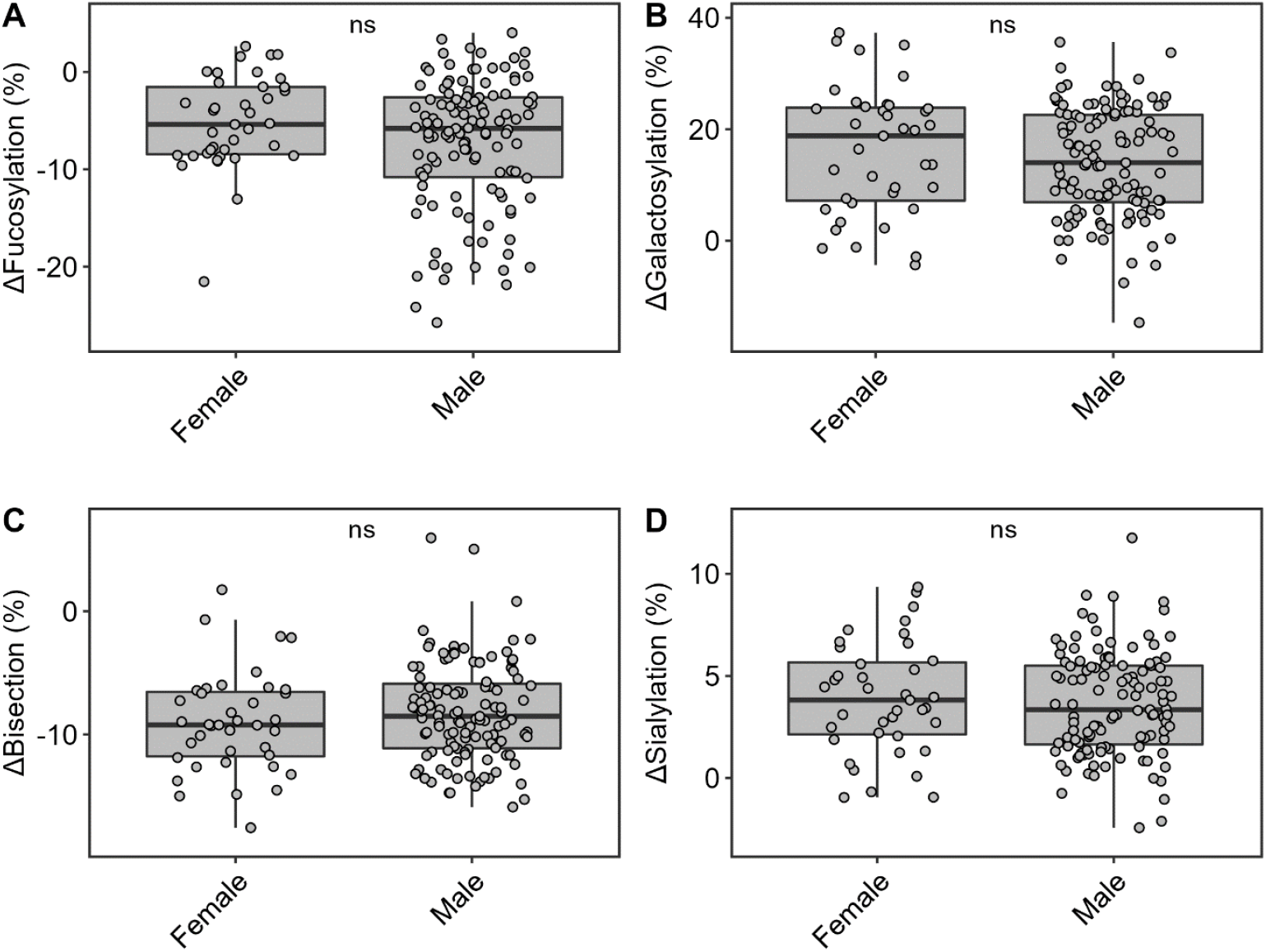
Comparison of Δglycosylation traits of female and male patients demonstrates the absence of a sex confounding effect. IgG1 **(A)** Δfucosylation, **(B)** Δgalactosylation, **(C)** Δbisection and **(D)** Δsialylation. Correction for the age and sex confounding effect was performed as described above **(Figure S2)**.

**Supplementary Figure 4.**
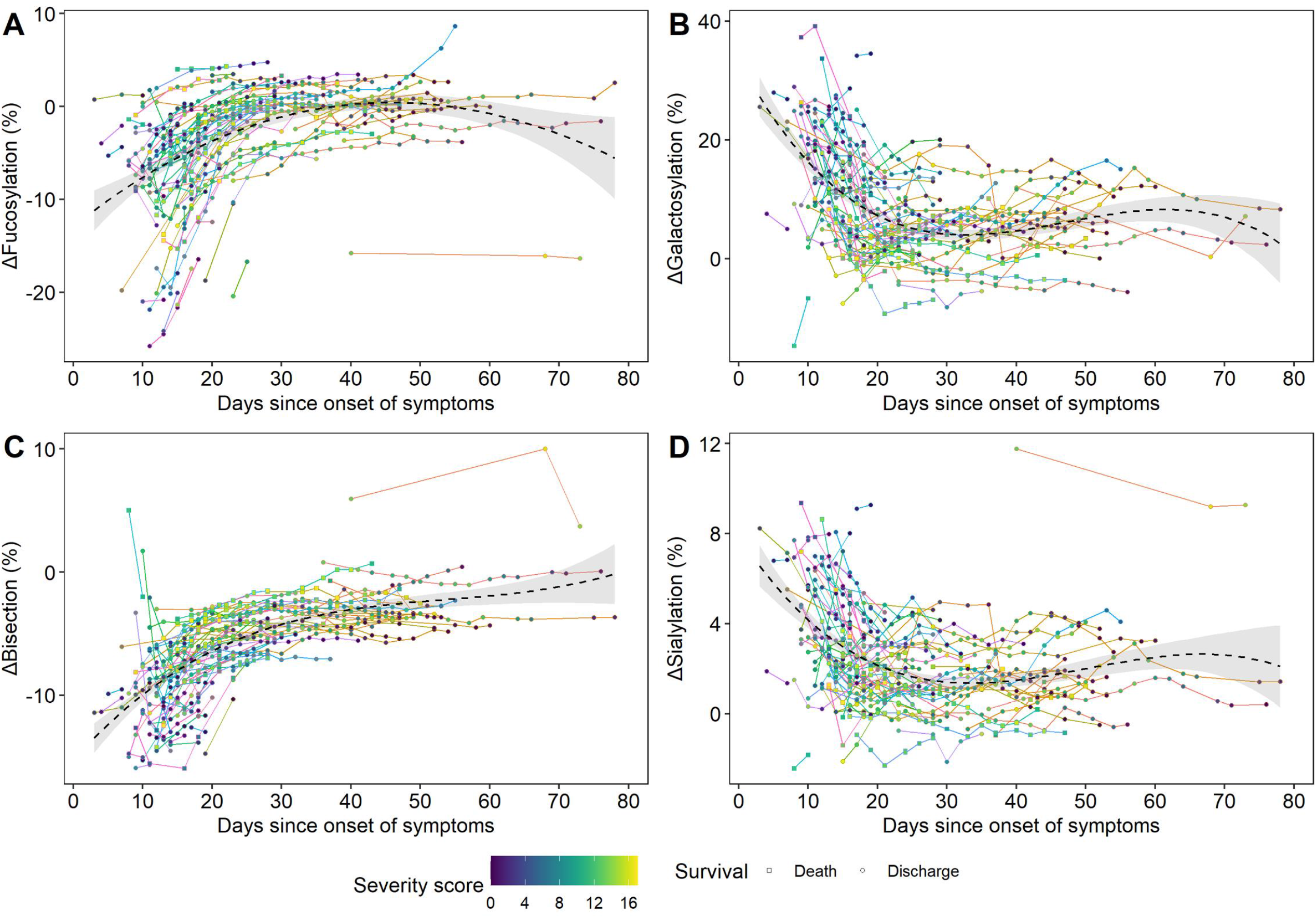
ΔIgG1 glycosylation dynamics during the entire hospitalization period. The time-course of Δglycosylation derived traits **(A)** fucosylation, **(B)** galactosylation, **(C)** bisection and **(D)** sialylation, as shown during hospitalization (n=111). Line colours correspond to a single COVID-19 patient, whilst the colour gradient in the circles/squares indicates the corresponding severity score (grey = NA). The shape displays whether a patient passed away (square) or was discharged alive (circle). The black dashed line with a grey 95% confidence interval band is a cubic polynomial fit over the shown datapoints to illustrate overall dynamics.

**Supplementary Figure 5.**
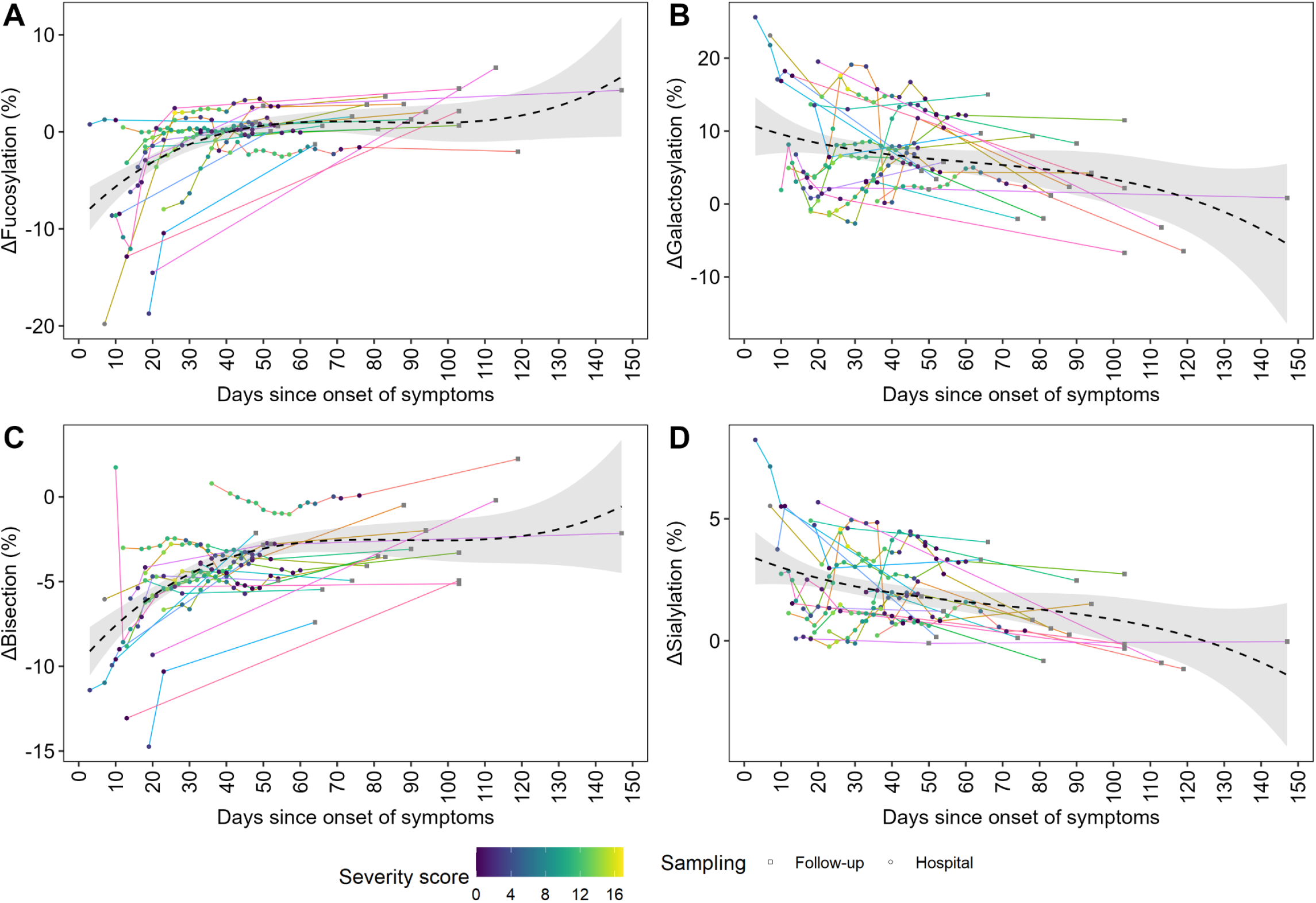
ΔIgG1 glycosylation dynamics of the subset of patients with a follow-up sample. The time-course of Δglycosylation derived traits **(A)** fucosylation, **(B)** galactosylation, **(C)** bisection and **(D)** sialylation, as shown during the hospitalization period and follow-up (n=19). Line colours correspond to a single COVID-19 patient, whilst the colour gradient in the circles/squares indicates the corresponding severity score (grey = NA). The circle and shape display whether the timepoint corresponds to a follow-up sample (square) or to a sample taken during hospitalization (circle). The black dashed line with a grey 95% confidence interval band is a cubic polynomial fit over the shown datapoints to illustrate overall dynamics.

**Supplementary Figure 6.**
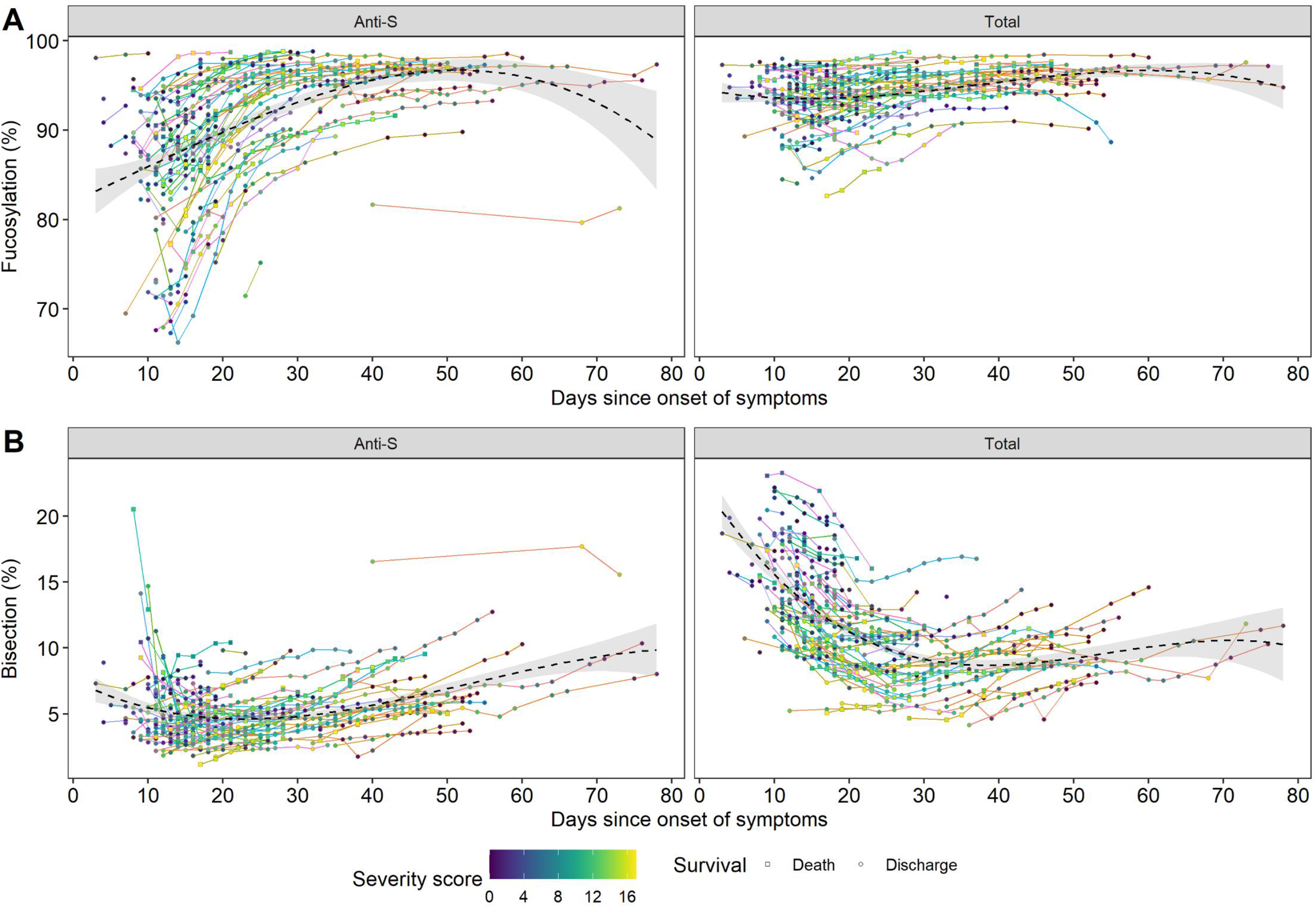
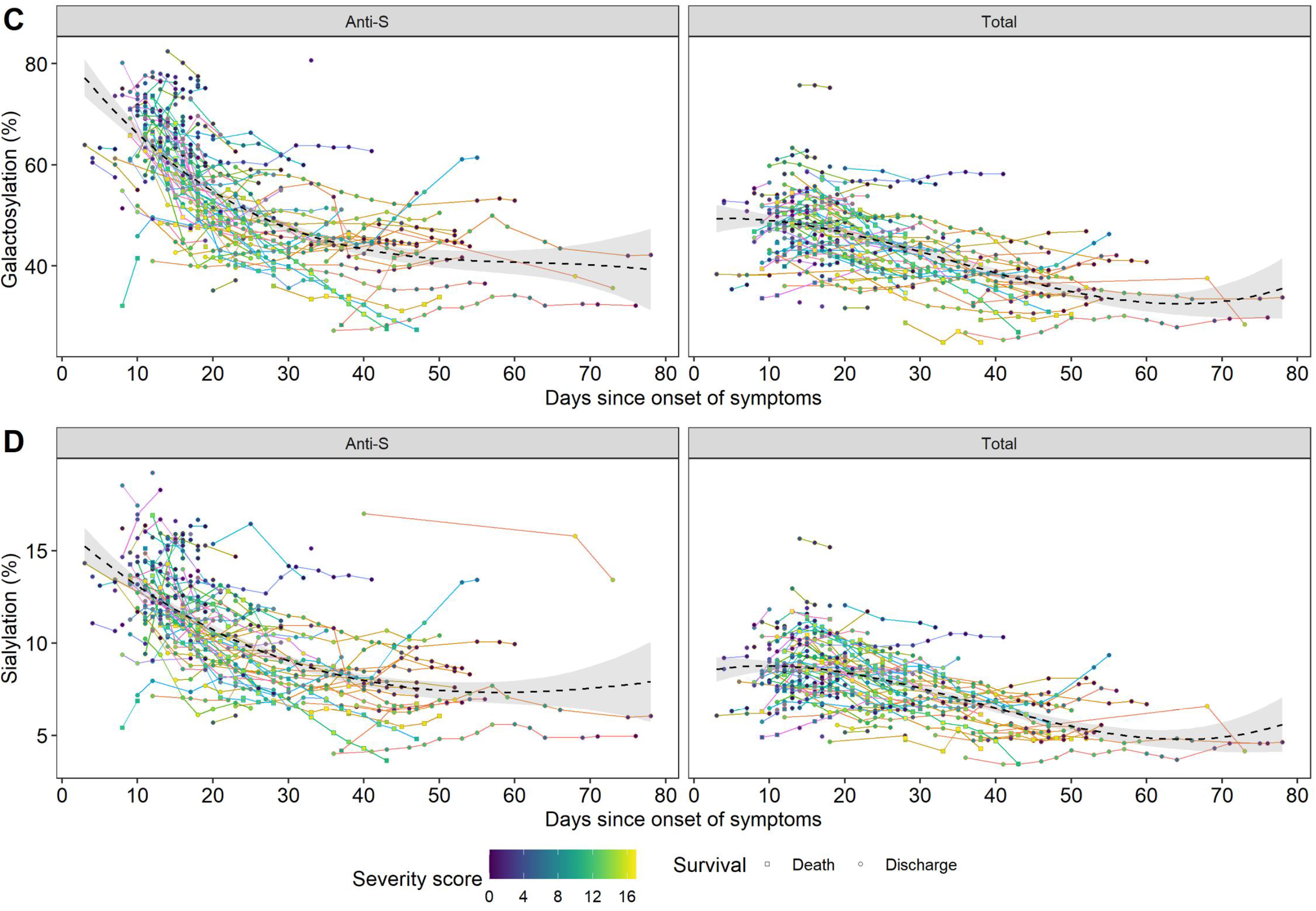
Anti-S and total IgG1 glycosylation dynamics during the entire hospitalization period. The time-course of glycosylation derived traits **(A)** fucosylation, **(B)** bisection, **(C)** galactosylation and **(D)** sialylation as shown during the hospitalization period (n=111). Anti-S IgG1 dynamics are shown in the left facets, whereas total IgG1 dynamics in the right facets in each panel. Line colors correspond to a single COVID-19 patient, whilst the colour gradient in the circles/squares indicates the corresponding severity score (grey = NA). The circle and shape display whether the patient passed away (square) or was discharged alive (circle) from the hospital. The black dashed line with a grey 95% confidence interval band is a cubic polynomial fit over the shown datapoints to illustrate overall dynamics. Note that the confounding effect of age largely influences the observed bisection **(Figure S2C)**, galactosylation **(Figure S2E)** and **(Figure S2G)** sialylation pattern, thereby has been corrected for age effects **(Figure S2)**.

**Supplementary Figure 7.**
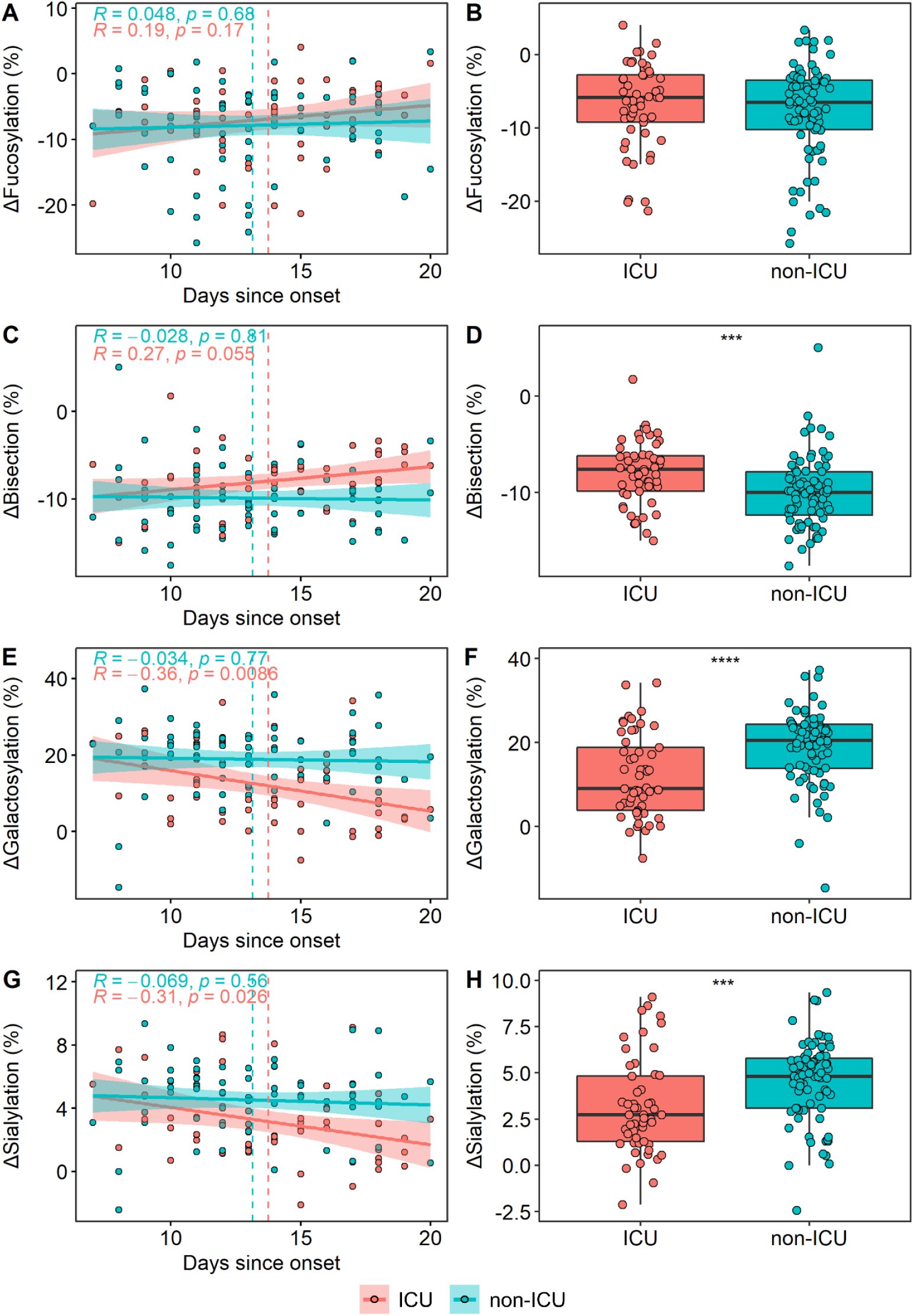
ICU (red) and non-ICU (blue) patients and corresponding ΔIgG1 glycosylation derived traits in a “days since onset of symptoms” subset of patients (n=129) to confirm that the observed differences (Figure 3) are not confounded by vast glycosylation dynamics. ΔIgG1 **(A)** fucosylation, **(C)** bisection, **(E)** galactosylation and **(G)** sialylation as a proxy of days since symptom onset. Shown in the inset are the Spearmen correlation coefficients (R) and *p*-values, respectively. The red (ICU) and blue (non-ICU) lines are linear regression lines and the corresponding band indicates the 95% confidence interval. The dashed vertical lines indicate group means. Comparison of corresponding ΔIgG1 **(B)** fucosylation, **(D)** bisection, **(F)** galactosylation and **(H)** sialylation levels between ICU and non-ICU patients. All datapoints correspond to baseline samples (time of hospitalization).

**Supplementary Figure 8.**
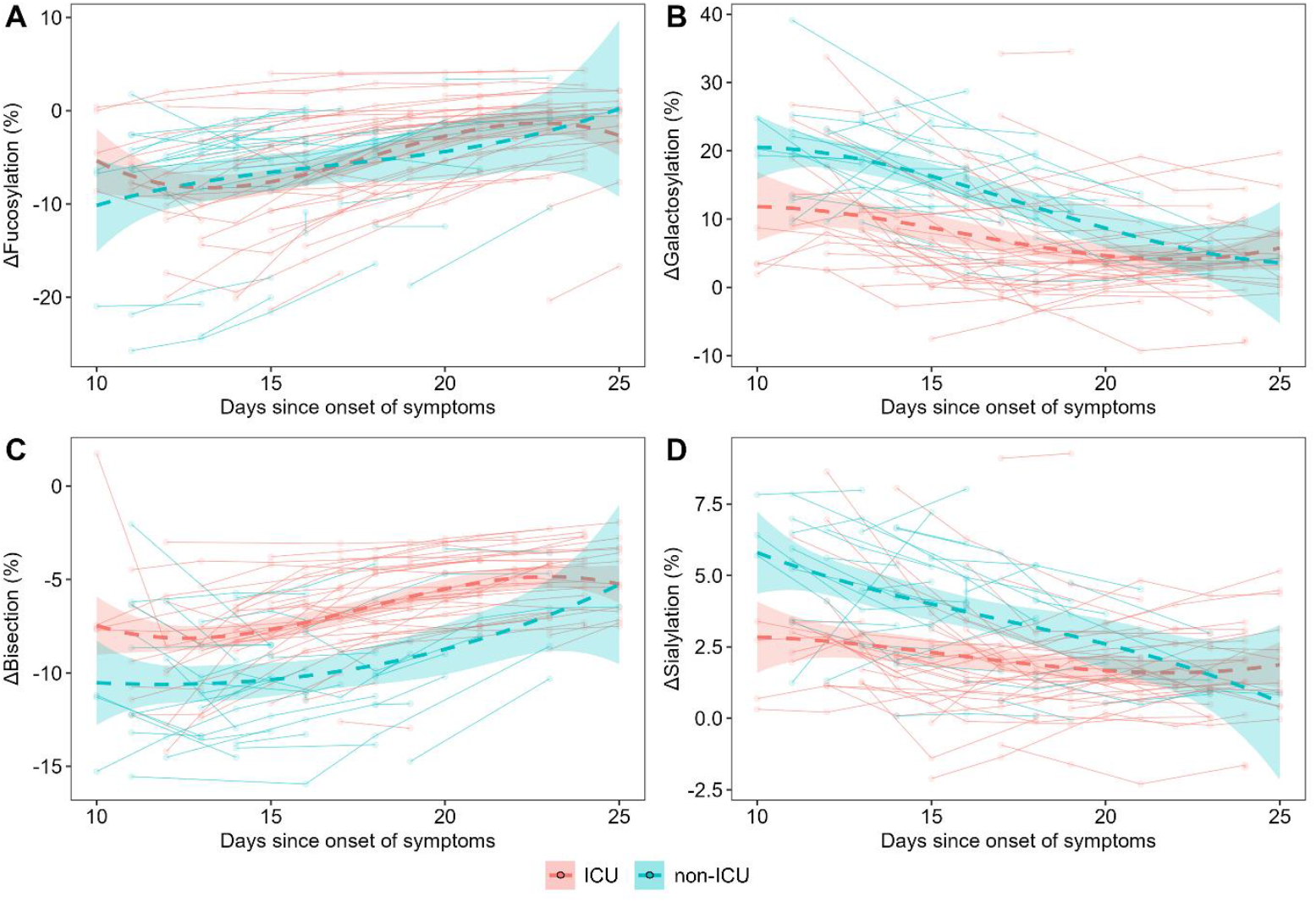
ΔIgG1 Δglycosylation dynamics of patients admitted to the ICU (n=48; red) and non-ICU (n=34; blue) treatments as shown between days 10 and 25. The time course of ΔIgG1 glycosylation traits **(A)** fucosylation, **(B)** galactosylation, **(C)** bisection and **(D)** sialylation. The dashed lines with 95% confidence interval bands are cubic polynomials fit over the shown datapoints to illustrate overall dynamics.

**Supplementary Figure 9.**
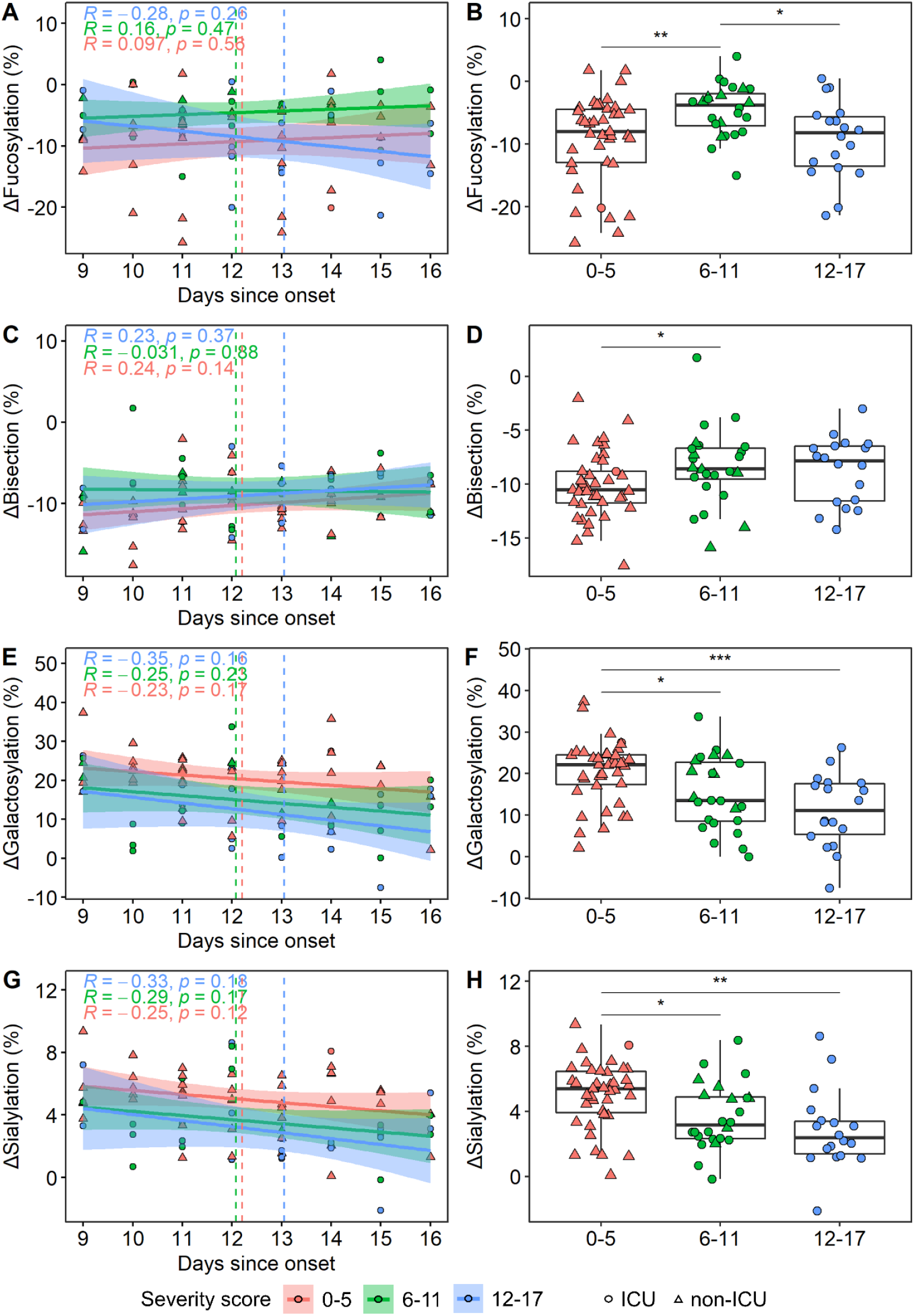
Patients in varying severity score groups 0-5 (red), 6-11 (green) and 12-17 (dark blue) and corresponding ΔIgG glycosylation derived traits in a “days since onset of symptoms” subset of patients to confirm that the observed differences (Figure 4) are not confounded by vast glycosylation dynamics. ΔIgG1 **(A)** fucosylation and **(C)** bisection, **(E)** galactosylation and **(G)** sialylation as a proxy of days since onset of subset of patients. Shown in the inset are the Spearmen correlation coefficients (R) and *p*-values, respectively. The red (0-5), green (6-11) and blue (12-17) lines are linear regression lines and the corresponding band indicates the 95% confidence interval. Dashed vertical lines indicate group mean. Comparison of corresponding ΔIgG1 **(B)** fucosylation and **(D)** bisection, **(F)** galactosylation and **(H)** sialylation levels between the three severity score groups. Circle indicates ICU patients, whereas squares indicate non-ICU patients. All datapoints correspond to baseline samples (time of hospitalization).

**Supplementary Figure 10.**
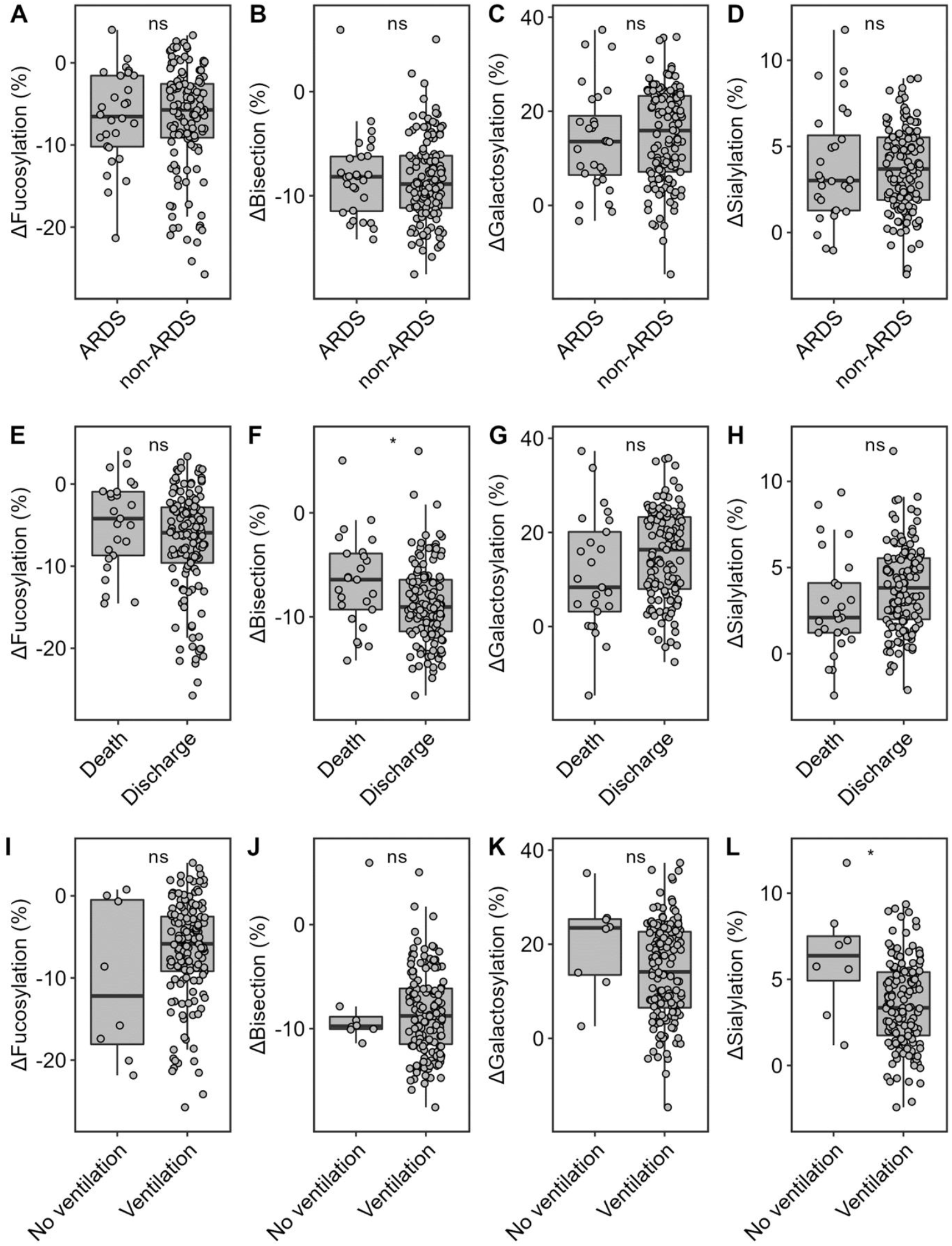
Comparison of acute respiratory distress syndrome (A-D), survival (E-H) and ventilation (I-L) subgroups of patients for glycosylation traits fucosylation (A, E, I), bisection (B, F, J), galactosylation (C, G, K) and sialylation (D, H, L). Bisection negatively associated with, survival, and sialylation negatively associated with ventilation. No other associations were found.

## References

1. Zhou P, Yang XL, Wang XG, et al. A pneumonia outbreak associated with a new coronavirus of probable bat origin. Nature 2020; 579(7798): 270–3.

2. Dong E, Du H, Gardner L. An interactive web-based dashboard to track COVID-19 in real time. The Lancet Infectious Diseases 2020; 20(5): 533–4.

3. Oran DP, Topol EJ. Prevalence of Asymptomatic SARS-CoV-2 Infection : A Narrative Review. Ann Intern Med 2020; 173(5): 362–7.

4. Long QX, Liu BZ, Deng HJ, et al. Antibody responses to SARS-CoV-2 in patients with COVID-19. Nat Med 2020; 26(6): 845–8.

5. Hu B, Guo H, Zhou P, Shi ZL. Characteristics of SARS-CoV-2 and COVID-19. Nat Rev Microbiol 2021; 19(3): 141–54.

6. Hoepel W, Chen HJ, Geyer CE, et al. High titers and low fucosylation of early human anti-SARS-CoV-2 IgG promote inflammation by alveolar macrophages. Sci Transl Med 2021; 13(596).

7. Ankerhold J, Giese S, Kolb P, et al. Circulating multimeric immune complexes drive immunopathology in COVID-19. bioRxiv 2021.

8. Bruhns P, Jonsson F. Mouse and human FcR effector functions. Immunol Rev 2015; 268(1): 25–51.

9. Lauc G, Pezer M, Rudan I, Campbell H. Mechanisms of disease: The human N-glycome. Biochim Biophys Acta 2016; 1860(8): 1574–82.

10. Dekkers G, Treffers L, Plomp R, et al. Decoding the Human Immunoglobulin G-Glycan Repertoire Reveals a Spectrum of Fc-Receptor-and Complement-Mediated-Effector Activities. Front Immunol 2017; 8: 877.

11. Ferrara C, Grau S, Jager C, et al. Unique carbohydrate-carbohydrate interactions are required for high affinity binding between FcgammaRIII and antibodies lacking core fucose. Proc Natl Acad Sci U S A 2011; 108(31): 12669–74.

12. van Osch TLJ, Nouta J, Derksen NIL, et al. Fc Galactosylation Promotes Hexamerization of Human IgG1, Leading to Enhanced Classical Complement Activation. J Immunol 2021; 207(6): 1545–54.

13. Larsen MD, de Graaf EL, Sonneveld ME, et al. Afucosylated IgG characterizes enveloped viral responses and correlates with COVID-19 severity. Science 2021; 371(6532).

14. Chakraborty S, Gonzalez J, Edwards K, et al. Proinflammatory IgG Fc structures in patients with severe COVID-19. Nat Immunol 2021; 22(1): 67–73.

15. Brouwer PJM, Caniels TG, van der Straten K, et al. Potent neutralizing antibodies from COVID-19 patients define multiple targets of vulnerability. Science 2020; 369(6504): 643–50.

16. Falck D, Jansen BC, de Haan N, Wuhrer M. High-Throughput Analysis of IgG Fc Glycopeptides by LC-MS. Methods Mol Biol 2017; 1503: 31–47.

17. Jansen BC, Falck D, de Haan N, et al. LaCyTools: A Targeted Liquid Chromatography-Mass Spectrometry Data Processing Package for Relative Quantitation of Glycopeptides. J Proteome Res 2016; 15(7): 2198–210.

18. Pucic M, Knezevic A, Vidic J, et al. High throughput isolation and glycosylation analysis of IgG-variability and heritability of the IgG glycome in three isolated human populations. Mol Cell Proteomics 2011; 10(10): M111 010090.

19. Clerc F, Reiding KR, Jansen BC, Kammeijer GS, Bondt A, Wuhrer M. Human plasma protein N-glycosylation. Glycoconj J 2016; 33(3): 309–43.

20. van Meijgaarden KE, Khatri B, Smith SG, et al. Cross-laboratory evaluation of multiplex bead assays including independent common reference standards for immunological monitoring of observational and interventional human studies. PLoS One 2018; 13(9): e0201205.

21. Escribano P, Alvarez-Uria A, Alonso R, et al. Detection of SARS-CoV-2 antibodies is insufficient for the diagnosis of active or cured COVID-19. Sci Rep 2020; 10(1): 19893.

22. Maine GN, Lao KM, Krishnan SM, et al. Longitudinal characterization of the IgM and IgG humoral response in symptomatic COVID-19 patients using the Abbott Architect. Journal of Clinical Virology 2020; 133.

23. Zlei M, Sidorov IA, Joosten S, et al. Absence of rapid T cell control corresponds with delayed viral clearance in hospitalised COVID-19 patients. ResearchSquare 2021.

24. Zhao J, Yuan Q, Wang H, et al. Antibody Responses to SARS-CoV-2 in Patients With Novel Coronavirus Disease 2019. Clin Infect Dis 2020; 71(16): 2027–34.

25. Beavis KG, Matushek SM, Abeleda APF, et al. Evaluation of the EUROIMMUN Anti-SARS-CoV-2 ELISA Assay for detection of IgA and IgG antibodies. J Clin Virol 2020; 129: 104468.

26. Knight SR, Ho A, Pius R, et al. Risk stratification of patients admitted to hospital with covid-19 using the ISARIC WHO Clinical Characterisation Protocol: development and validation of the 4C Mortality Score. BMJ 2020; 370: m3339.

27. Kristic J, Vuckovic F, Menni C, et al. Glycans are a novel biomarker of chronological and biological ages. J Gerontol A Biol Sci Med Sci 2014; 69(7): 779–89.

28. Bye AP, Hoepel W, Mitchell JL, et al. Aberrant glycosylation of anti-SARS-CoV-2 IgG is a pro-thrombotic stimulus for platelets. Blood 2021; 138(16): 1481–9.

29. Petrovic T, Alves I, Bugada D, et al. Composition of the immunoglobulin G glycome associates with the severity of COVID-19. Glycobiology 2021; 31(4): 372–7.

30. Kox M, Waalders NJB, Kooistra EJ, Gerretsen J, Pickkers P. Cytokine Levels in Critically Ill Patients With COVID-19 and Other Conditions. JAMA 2020; 324(15): 1565–7.

31. Ackerman ME, Crispin M, Yu X, et al. Natural variation in Fc glycosylation of HIV-specific antibodies impacts antiviral activity. J Clin Invest 2013; 123(5): 2183–92.

32. Wang TT, Sewatanon J, Memoli MJ, et al. IgG antibodies to dengue enhanced for FcgammaRIIIA binding determine disease severity. Science 2017; 355(6323): 395–8.

33. Kapur R, Della Valle L, Sonneveld M, et al. Low anti-RhD IgG-Fc-fucosylation in pregnancy: a new variable predicting severity in haemolytic disease of the fetus and newborn. Br J Haematol 2014; 166(6): 936–45.

34. Kapur R, Kustiawan I, Vestrheim A, et al. A prominent lack of IgG1-Fc fucosylation of platelet alloantibodies in pregnancy. Blood 2014; 123(4): 471–80.

35. Sonneveld ME, de Haas M, Koeleman C, et al. Patients with IgG1-anti-red blood cell autoantibodies show aberrant Fc-glycosylation. Sci Rep 2017; 7(1): 8187.

36. Sonneveld ME, Koelewijn J, de Haas M, et al. Antigen specificity determines anti-red blood cell IgG-Fc alloantibody glycosylation and thereby severity of haemolytic disease of the fetus and newborn. Br J Haematol 2017; 176(4): 651–60.

37. Sonneveld ME, Natunen S, Sainio S, et al. Glycosylation pattern of anti-platelet IgG is stable during pregnancy and predicts clinical outcome in alloimmune thrombocytopenia. Br J Haematol 2016; 174(2): 310–20.

38. de Haan N, Boeddha NP, Ekinci E, et al. Differences in IgG Fc Glycosylation Are Associated with Outcome of Pediatric Meningococcal Sepsis. mBio 2018; 9(3).

39. Thomann M, Schlothauer T, Dashivets T, et al. In vitro glycoengineering of IgG1 and its effect on Fc receptor binding and ADCC activity. PLoS One 2015; 10(8): e0134949.

40. Lu LL, Das J, Grace PS, Fortune SM, Restrepo BI, Alter G. Antibody Fc Glycosylation Discriminates Between Latent and Active Tuberculosis. J Infect Dis 2020; 222(12): 2093–102.

41. Peschke B, Keller CW, Weber P, Quast I, Lunemann JD. Fc-Galactosylation of Human Immunoglobulin Gamma Isotypes Improves C1q Binding and Enhances Complement-Dependent Cytotoxicity. Front Immunol 2017; 8: 646.

42. Carvelli J, Demaria O, Vely F, et al. Association of COVID-19 inflammation with activation of the C5a-C5aR1 axis. Nature 2020; 588(7836): 146–50.

43. Kaneko Y, Nimmerjahn F, Ravetch JV. Anti-inflammatory activity of immunoglobulin G resulting from Fc sialylation. Science 2006; 313(5787): 670–3.

44. Washburn N, Schwab I, Ortiz D, et al. Controlled tetra-Fc sialylation of IVIg results in a drug candidate with consistent enhanced anti-inflammatory activity. Proc Natl Acad Sci U S A 2015; 112(31): E4339.

45. Quast I, Keller CW, Maurer MA, et al. Sialylation of IgG Fc domain impairs complement-dependent cytotoxicity. J Clin Invest 2015; 125(11): 4160–70.

## References

1. Gudelj I, Lauc G, Pezer M. Immunoglobulin G glycosylation in aging and diseases. Cell Immunol 2018; 333: 65–79.

